# Genome-wide analysis of blood lipid metabolites in over 5,000 South Asians reveals biological insights at cardiometabolic disease loci

**DOI:** 10.1101/2020.10.16.20213520

**Authors:** Eric L. Harshfield, Eric B. Fauman, David Stacey, Dirk S. Paul, Daniel Ziemek, Rachel M. Y. Ong, John Danesh, Adam S. Butterworth, Asif Rasheed, Taniya Sattar, Zameer-ul-Asar, Imran Saleem, Zoubia Hina, Unzila Ishtiaq, Nadeem Qamar, Nadeem Hayat Mallick, Zia Yaqub, Tahir Saghir, Syed Nadeem Hasan Rizvi, Anis Memon, Mohammad Ishaq, Syed Zahed Rasheed, Fazal-ur-Rehman Memon, Anjum Jalal, Shahid Abbas, Philippe Frossard, Danish Saleheen, Angela M. Wood, Julian L. Griffin, Albert Koulman

## Abstract

**Background:** Genetic, lifestyle, and environmental factors can lead to perturbations in circulating lipid levels and increase risk of cardiovascular and metabolic diseases. However, how changes in individual lipid species contribute to disease risk is often unclear. Moreover, little is known about the role of lipids on cardiovascular disease in Pakistan, a population historically underrepresented in cardiovascular studies.

**Methods:** We characterised the genetic architecture of the human blood lipidome in 5,662 hospital controls from the Pakistan Risk of Myocardial Infarction Study (PROMIS) and 13,814 healthy British blood donors from the INTERVAL study. We applied a candidate causal gene prioritisation tool to link the genetic variants associated with each lipid to the most likely causal genes, and Gaussian Graphical Modelling network analysis to identify and illustrate relationships between lipids and genetic loci.

**Results:** We identified 359 genetic associations with 255 lipids measured using direct infusion high-resolution mass spectrometry in PROMIS, and 616 genetic associations with 326 lipids in INTERVAL. Our analyses revealed new biological insights at genetic loci associated with cardiometabolic diseases, including novel lipid associations at the *LPL, MBOAT7, LIPC, APOE-C1-C2-C4, SGPP1*, and *SPTLC3* loci.

**Conclusions:** Our findings, generated using a distinctive lipidomics platform in an understudied South Asian population, strengthen and expand the knowledge base of the genetic determinants of lipids and their association with cardiometabolic disease-related loci.

## BACKGROUND

Mass spectrometry-based lipidomics, which aims to capture information on the full complement of lipid metabolites in a given biological sample [1], holds the potential to identify novel insights leading to lipid regulation and dyslipidaemia, potentially providing new mechanisms that link lipid perturbances with cardiometabolic disorders. While pathways underlying dyslipidaemia have been widely studied, we still do not understand how individual lipid species are regulated or contribute to disease. With increasing rates of cardiometabolic diseases in low- and middle-income countries, there is a need for well-powered studies to understand the mechanisms that lead to such disorders in these settings. This need is especially acute for genetic studies where the overrepresentation of individuals of European ancestry amongst genotyped cohorts has led to ancestral bias in effect size estimates at both the genotype and polygenic score levels [2].

In this study, we aimed to identify novel genetic associations with lipid metabolites in an understudied South Asian population and determine plausible metabolic pathways for the significantly associated lipid metabolites. We performed a comprehensive interrogation of genetic influences on the human blood serum lipidome using direct infusion high resolution mass spectrometry (DIHRMS). We quantified 360 lipid metabolites in 5,662 individuals from Pakistan, from which we identified 359 genotype–lipid associations (lipid quantitative trait loci, or lipid QTLs [3, 4]) at 24 independent loci, providing new insights into lipid metabolism and its impact on cardiovascular and metabolic diseases.

To help disentangle which of these findings are specific to the Pakistani population and which are unique to the lipid platform itself, we also carried out a parallel set of analyses using the same lipidomics platform in a much larger cohort of individuals from the UK. We measured 432 lipid metabolites in 13,814 healthy British blood donors, from which we identified 616 lipid QTLs at 38 independent loci.

## METHODS

### Study description

Our primary analyses involved a subset of participants from the Pakistan Risk of Myocardial Infarction Study (PROMIS), a case-control study of first-ever acute myocardial infraction (MI) in nine urban centres in Pakistan consisting of approximately 16,700 cases and 18,600 controls. Details of PROMIS have been described previously [5]. In this analysis we analysed controls (individuals free from MI at baseline), who were identified and recruited at the same hospitals as cases according to the following order of priority: (1) visitors of patients attending the outpatient department, (2) patients attending outpatient clinics for non-cardiac-related symptoms, and (3) non-first-degree relative visitors of MI cases. The present analysis involved serum samples from 5,662 PROMIS controls for which genetic and lipid-profiling data were available. Ethical approval was obtained from the relevant ethics committee of each of the institutions involved in participant recruitment and the Center for Non-Communicable Diseases in Karachi, Pakistan, and informed consent was obtained from each participant recruited into the study, including for use of samples in genetic, biochemical, and other analyses.

Comparative, parallel analyses were performed in INTERVAL, a prospective cohort study of approximately 50,000 healthy blood donors from the UK. The present analyses involved 13,814 participants from INTERVAL with both genetic and lipid-profiling data. Details concerning the INTERVAL study, including DNA extraction and genotyping, lipid profiling, and genome-wide association analyses are provided in the Supplementary Methods (Additional file 1).

### Lipid profiling

Lipid levels in human serum were quantified using direct infusion high-resolution mass spectrometry (DIHRMS) using an Exactive Orbitrap (Thermo, Hemel Hampstead, UK). Data processing, peak-picking, normalisation, cleaning, and quality control were performed to identify and record signals for 360 known lipids in 5,662 PROMIS participants. The 360 lipids corresponded to five broad lipid categories (fatty acyls and derivatives, glycerolipids, glycerophospholipids, sphingolipids, and sterol lipids), which are further subdivided into fourteen lipid subclasses (Supplementary Table 1 in Additional file 2). We have previously described all the details of our lipid profiling, data processing, quality control, and peak-picking process [6]. In brief, lipid profiles were obtained using an open-profiling technique that measured all lipid species across a spectrum. We developed a novel peak-picking algorithm [6] to select all lipids within an *m/z* window of 185-1000, with a time window of 20-70 seconds for lipids in positive ionisation mode and 95-145 seconds for lipids in negative ionisation mode. A lipid list containing all known lipids within this *m/z* range was used to extract information on the lipid concentrations at specific peaks of interest, consisting of 1,305 lipids in positive ionisation mode and 3,772 lipids in negative ionisation mode. Quality control samples and blanks were used to remove lipids that were not able to be detected or had poor quality of assessment, resulting in a final list of 360 distinct lipid annotations across both ionisation modes.

### Genotyping and imputation

DNA from PROMIS participants was extracted from leukocytes in Pakistan and genotyped at the Wellcome Sanger Institute in Cambridge, UK on either (1) the Illumina 660-Quad GWAS platform, which consisted of 527,925 genotyped autosomal variants after quality control (QC) steps were performed, or (2) the Illumina HumanOmniExpress GWAS platform, which consisted of 643,333 genotyped autosomal variants after QC. Genetic samples were removed if (1) they were heterozygosity outliers (heterozygosity > mean ± 3 SD), (2) the sample call rate was less than 97%, (3) there was discordant sex between genetically-inferred and self-reported sex, or (4) they were duplicate or related pairs (kinship coefficient > 0.375). Single nucleotide polymorphisms (SNPs) were excluded if (1) the SNP call rate was less than 97%, (2) there was evidence of departure from Hardy-Weinberg Equilibrium (HWE) at a *P*-value of less than 1 x 10^−7^, or (3) the minor allele frequency (MAF) was less than 1%. Imputation was applied to the cleaned PROMIS datasets using the 1000 Genomes Project March 2012 (v3) release [7] as the reference panel. Imputation was conducted using IMPUTE v2.1.0 [8] using 5-Mb non-overlapping intervals for the whole genome. Once imputation had been performed for the samples on both genotyping platforms separately, there were over 7.2 million imputed SNPs available for analyses in either dataset before further QC. SNPs were removed if they were poorly imputed, i.e. if they had an information score (an assessment of the level of accuracy of imputation) < 80%. The results were then extracted from the output files, and once the final QC filters were reapplied, 6,720,657 SNPs were available for analyses of the lipidomics data. In total, 5,662 individuals from PROMIS had concomitant information on lipidomics data and imputed SNPs.

### Primary genome-wide association analyses

In PROMIS, linear regression was used to determine the association of each lipid with each SNP using SNPTEST v2.4.1 [9], which was performed separately for the samples genotyped on each of the two genetic platforms. Residuals were calculated from the null model for each lipid, which included adjustment for age group, sex, date of survey, plate (batch), and fasting status. To account for population stratification and genetic substructure in the data, principal component analysis was conducted on the multi-dimensional scaling matrix created from autosomal SNPs as implemented in PLINK; the first six principal components were subsequently added to each model. A missing data likelihood score test was used when testing for association at imputed SNPs to account for genotype uncertainty. Beta estimates and standard errors from the association results for the two genetic platforms were combined in a fixed-effect inverse-variance-weighted meta-analysis using METAL version 2011-03-25 [10]. The threshold for genome-wide significance level was set to *P* < 8.929 x 10^−10^, which corrected for multiple testing by dividing the standard genome-wide significance level (5 x 10^−8^) by the number of principal components (56) that explained over 95% of the variance in the levels of the lipids. All traits gave genomic inflation factors (*λ*) in the meta-analysis less than 1.05 [mean (SD) 1.0139 (0.0129); range 0.9741-1.0455], indicating that there was little evidence of systematic bias in the test statistics.

To verify the robustness and validity of the results, post-analysis quality control (QC) was performed by comparing the meta-analysis results with the results on each GWAS platform. The lead SNPs from the meta-analysis were only kept if they (1) passed QC in the raw SNPTEST results from both GWAS platforms (i.e. HWE *P* < 1 x 10^−7^, call rate < 0.97, MAF < 0.01, and info score < 0.80); (2) had beta (*β*) estimates in the same direction on both platforms (i.e. betas were both negative or both positive); and (3) had *P* < 0.01 on both platforms (with *P* < 8.9 x 10^−10^ in the meta-analysis).

### Genome-wide analysis of ratios of lipids

A second discovery step was carried out in PROMIS by testing genome-wide associations on 26 pairwise ratios of lipid concentrations. Ratios were identified based on those that had strong biological rationales and that acted through thoroughly understood metabolic pathways (Supplementary Table 5). Meta-analysis was performed to combine results from the two genotyping platforms using a fixed-effect inverse-variance weighted meta-analysis. Since there were fewer statistical tests for the ratios than for the individual lipids, the combined results file for each ratio was filtered using the standard threshold for genome-wide significance of *P* < 5 x 10^−8^.

### Conditional analyses

We conducted conditional analyses on the significant loci from the meta-analysis results of the univariate GWAS for each lipid in PROMIS. All SNPs were selected where *P* < 8.9 x 10^−10^, the 5-Mb chunks were identified where each of these SNPs were located, and the lead SNPs were selected within each chunk that had the strongest *P*-value. On an individual lipid basis, for each 5-Mb chunk that was identified, SNPTEST was run on the imputed data for each genotyping platform using the same null model as before, except also conditioning on the lead SNP in the identified chunk. The results from the samples analysed on each genotyping platform were combined in a meta-analysis using METAL as described above, and any SNPs where *P* < 8.9 x 10^−10^ were identified. The lead SNP from the meta-analysed results of the first conditional analysis (i.e. the SNP with the strongest *P*-value) was identified, and this process was repeated for each chunk. Additional SNPs to be conditioned on were repeatedly added to the model on each chunk for each lipid until there were no more significant SNPs left within that chunk. The final set of SNPs that were “conditionally independent” for each lipid were combined into a single list across all lipids, resulting in 359 SNP-lipid associations (lipid QTLs) for 255 lipids, or 90 unique lead SNPs. These variants were grouped into 24 loci using a distance measure of ±500-Kb.

We identified the proportion of variation in the lipidome explained by inherited genetic variants by regressing each lipid on the number of copies of each allele held by each participant for each of the conditional analysis sentinel SNPs.

### Candidate gene annotation

In order to prioritise candidate genes that might underpin the genotype—lipid associations, we applied the ProGeM framework (Supplementary Figure 5 in Additional file 1) to both PROMIS and INTERVAL [11]. In addition to reporting the nearest gene to the sentinel variant, ProGeM combines information from complementary “bottom-up” and “top-down” approaches to assess the credibility of potential candidate genes [11] (Supplementary Table 7). In the bottom-up approach, we annotated SNPs according to their putative effects on proximal gene function by examining whether these SNPs influence protein sequencing, gene splicing, and/or mRNA levels of a local gene (Supplementary Table 8). Conversely, in the top-down approach, we annotated SNPs according to previous knowledge concerning local gene function by examining whether proximal genes have been previously implicated in lipid metabolism (Supplementary Table 9). In cases where (1) SNPs were purported to exert effects on more than one local gene and/or (2) more than one local gene was previously implicated in lipid metabolism, we assigned SNPs to multiple genes rather than force-assigning each to a single gene. In cases where it was not possible to annotate SNPs using either the bottom-up or top-down approach, we assigned the SNPs to their nearest gene. Further details of the candidate gene annotation approach that we followed are described in the Supplementary Methods (Additional file 1).

After performing comprehensive annotation of SNPs as per the bottom-up and top-down procedures, we then integrated this information to try to predict the most likely causal gene(s) using a hierarchical approach as follows: (1) For those lead SNPs where the same gene was highlighted by both the bottom-up and the top-down approach, we selected this gene as the putative causal gene; (2) If both the SNP (from this study) and the proximal gene (from IPA) were associated with the same lipid subclass, we made further SNP-gene assignments accordingly; (3) Finally, for each of the remaining lead SNPs, we assigned the highest scoring top-down gene and any bottom-up genes as the likely causal gene(s).

Separately, we assigned an expertly-curated causal gene to each variant and compared the predicted causal genes identified by the functional annotation pipeline to assess concordance and validate the pipeline.

### Gaussian Graphical Modelling

As described previously [6], we estimated a Gaussian Graphical Model (GGM) on the normalised relative intensities of the lipids in PROMIS to better resolve lipid cross-correlations. The GGM resulted in a set of edges in which each edge connected two detected lipids if their cross-correlation conditioned on all other lipids was significantly different from zero. Subjects with more than 10% missing lipids as well as lipids with more than 20% missing subjects were removed from the analysis. The “genenet” R package was used to infer the GGM [12]. A similar approach for metabolomics data has been suggested previously [13]. To focus on strong effects we retained only edges in the model that met an FDR cutoff of 0.05 and had a partial correlation coefficient greater than 0.2.

### Fatty acid chain enrichment analysis

We manually annotated detected lipids in PROMIS with their constituent fatty acid chains. For each combination of fatty acid chains, we counted the number of GGM edges connecting lipids with that specific combination, which we used to directly estimate *P*-values of enrichment and depletion. To test whether edges from the GGM were enriched for any combination of fatty acid chains, we permuted the annotation 1000 times using the R package “BiRewire” [14], keeping the number of annotations per lipid and fatty acid chain constant.

### Network of genetic and metabolic associations

We used Cytoscape v3.2.1 [15] to generate a network of associations between genes and lipid subclasses in PROMIS (Figure 3). Using a previously described approach [16], we constructed a GGM to connect lipids to each other based on partial correlation coefficients, and we also connected lipids with genetic loci using the conditional analysis results, with one link for each genome-wide significant association. The full network facilitates visualisation of the genetic determinants of human metabolism and the relationships between genetic loci and lipid subclasses.

**Figure 1.**
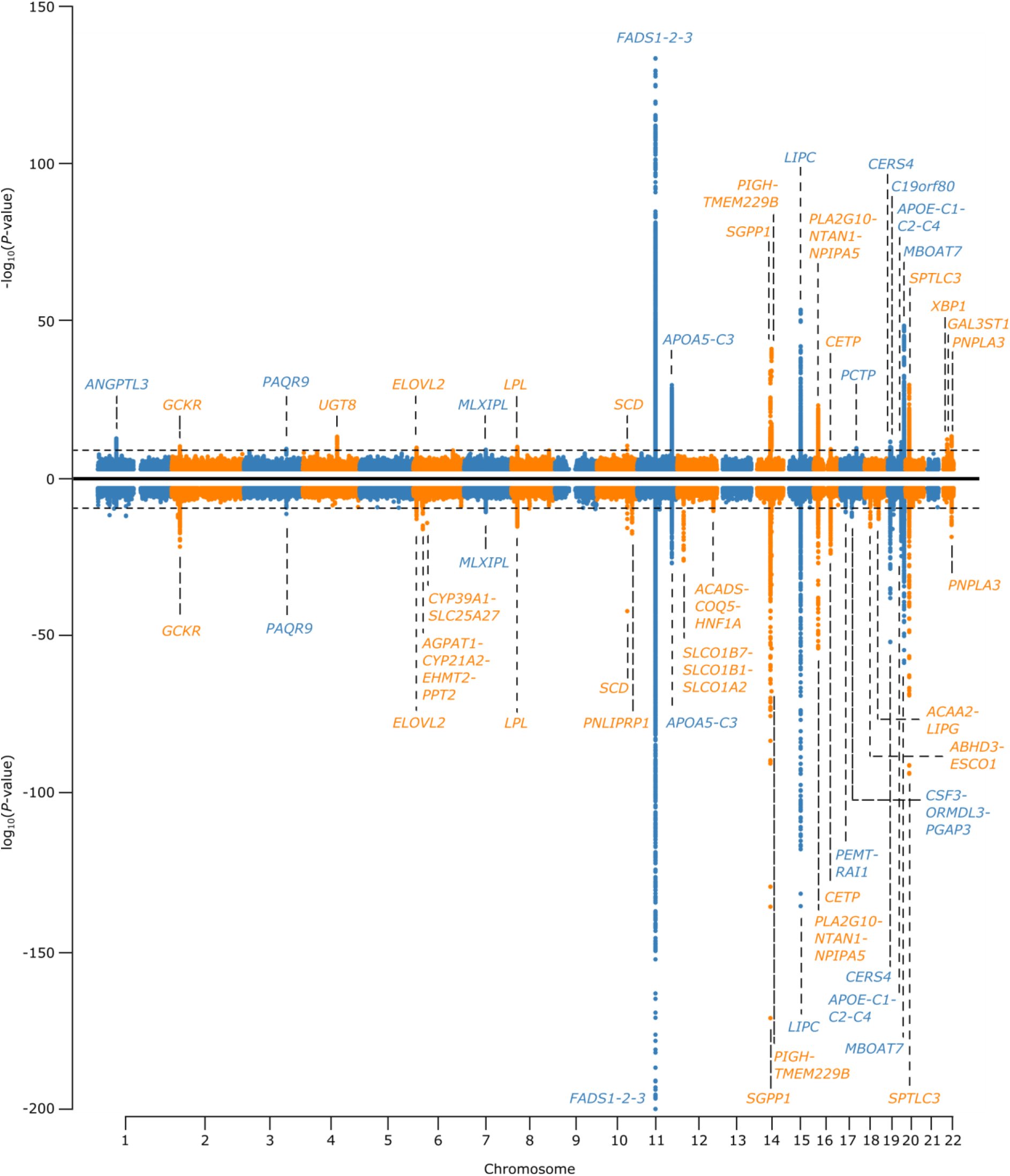
Miami plot of combined association results from genome-wide association analysis for all lipids in PROMIS and INTERVAL. The combined association results are shown for all lipids with each variant in PROMIS (top) and INTERVAL (bottom). *P*-values > 1 x 10^−3^ have been truncated at 1 x 10^−3^, and *P*-values < 1 x 10^−200^ have been truncated at 1 x 10^−200^. Actual *P*-value for lead SNP in *FADS-1-2-3* locus in INTERVAL is 1.6 x 10^−286^.

**Figure 2.**
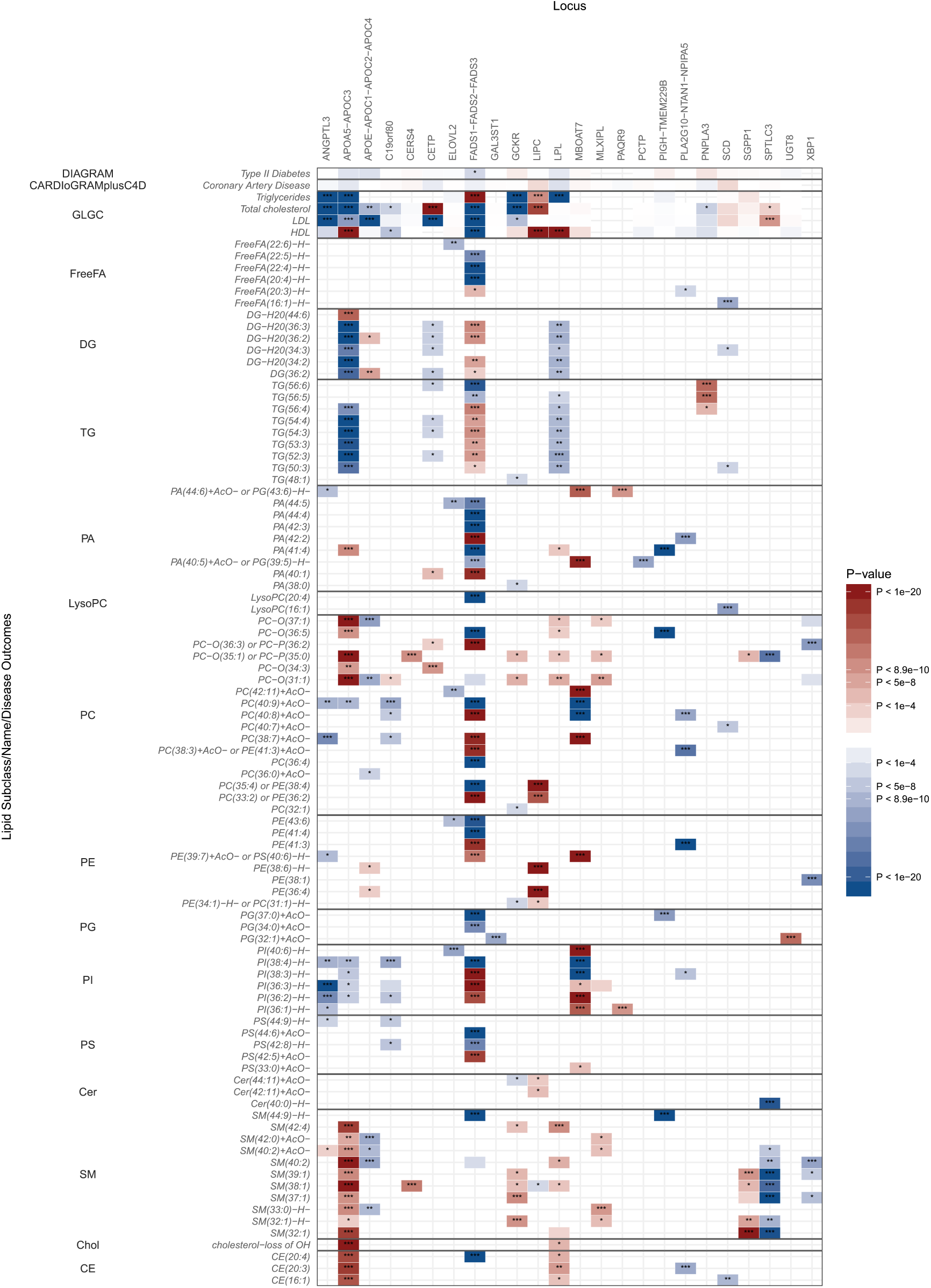
Heat map showing associations of significant loci from conditional analyses with selected lipid metabolites in PROMIS. The effect estimates of the associations between significant variants and selected lipids are plotted as a heat map. Results are shown for selected top lipids with the strongest associations within each subclass (rows) against the most strongly associated genetic variant within each locus (columns). The associations with major lipids from the GLGC (total cholesterol, HDL-C, LDL-C, and triglycerides), DIAGRAM Consortium (type 2 diabetes), and CARDIoGRAMplusC4D Consortium (coronary artery disease) are also shown. The magnitude and direction of the effect estimates (standardised per 1-SD) are indicated by a colour scale, with blue indicating a negative association and red indicating a positive association with respect to the SNP effect on the trait. Asterisks indicates the degree of significance of the *P*-values of association. * = *P* < 1 x 10^−4^; ** = *P* < 5 x 10^−8^; *** = *P* < 8.9 x 10^−10^.

**Figure 3.**
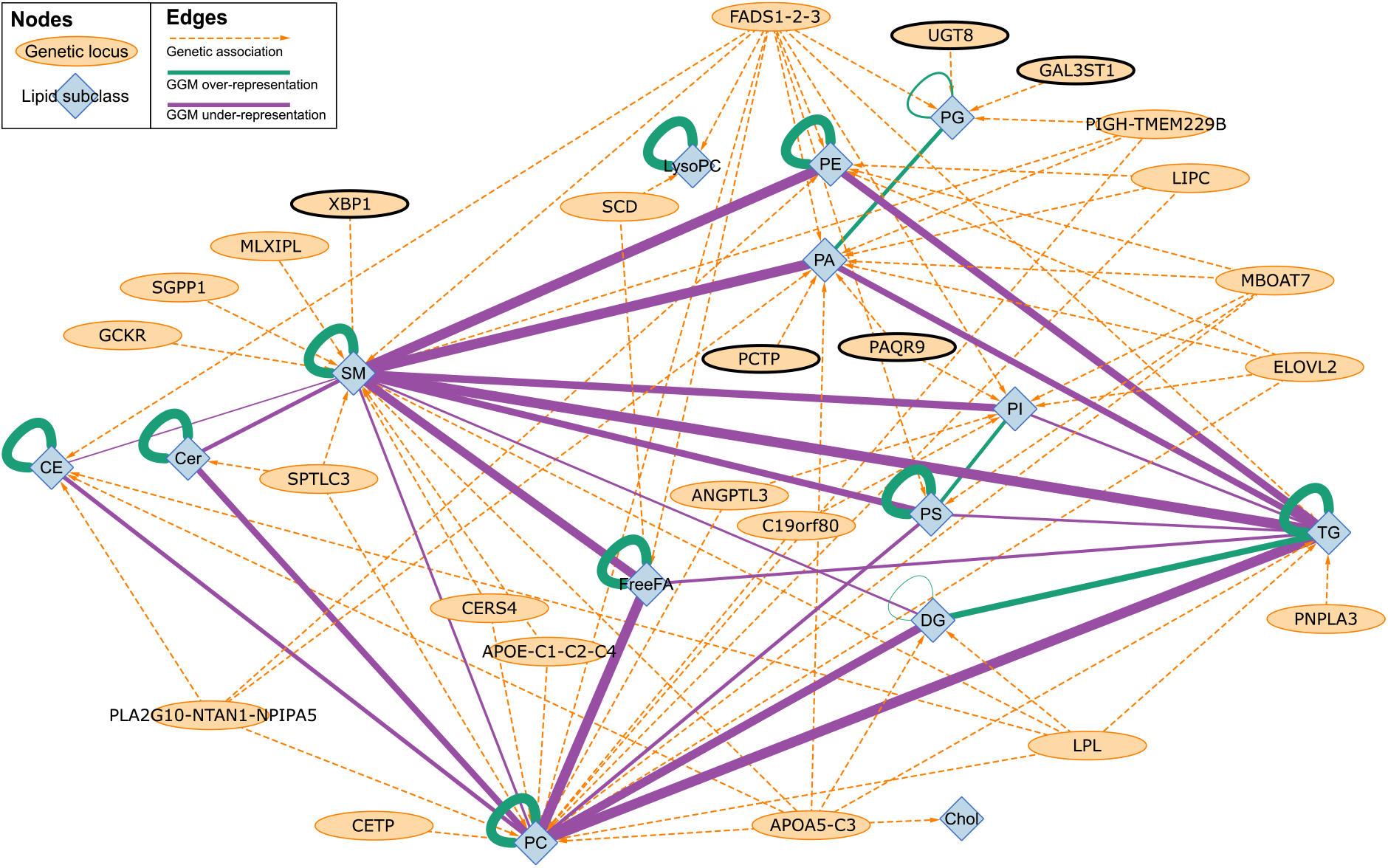
Combined network graph summarising genetic associations and a Gaussian graphical model (GGM) relating to levels of individual lipid species in PROMIS. Nodes representing genetic loci are each labelled with the most likely “causal” gene at that locus according to our functional annotation (see Methods). In order for an edge to be drawn between a genetic locus and a lipid subclass, there must have been a minimum of one variant at that locus significantly (*P* < 8.9 x 10^−10^) associated with a minimum of one lipid species belonging to that lipid subclass. Edges between lipid subclasses indicate whether there was either a significant over- (green) or under- (purple) representation (the magnitude is indicated in the thickness of the edges) of GGM connections between lipid species belonging to different lipid subclasses.

The network diagrams were created by combining two parts to integrate different sources of information. The first part was created by loading the reported associations between lipids and genes into Cytoscape. Lipid species were clustered according to the lipid subclass they belong to, resulting in fourteen distinct lipid subclass nodes in the network. The 90 identified lead SNPs from the conditional analyses were clustered according to their corresponding predicted causal gene(s), which was determined using the ProGeM framework [11]. In cases where it was not possible to confidently identify a single predicted causal gene, loci were entered into the network instead. For the second part, a functional interaction network consisting solely of our list of predicted causal genes/loci was created in Cytoscape using interaction network data downloaded from Ingenuity Pathway Analysis (IPA) that had been merged using in-house R scripts to create a .sif file.

For loci with multiple potential causal genes, interaction networks for all genes were extracted from IPA and an edge was drawn if at least one gene at that locus functionally interacts with another of our lipid-associated genes according to IPA. Finally, these two parts were merged together by node names (i.e. gene symbols). No enrichment statistics (e.g. KEGG pathways or GO terms) or other statistical information was used to produce the network, since this information was already incorporated to inform the predictions of the most likely “causal” genes, and would therefore invalidate the conclusions if it was also used to inform the network.

A second network diagram was created containing a subset of the first network containing only the triglyceride species (Figure 4). It also provides more detail as it shows the individual triglycerides rather than the lipid subclass as a whole. Thus, it portrays the partial correlations of the triglycerides with each other and the association of each triglyceride with genetic loci.

**Figure 4.**
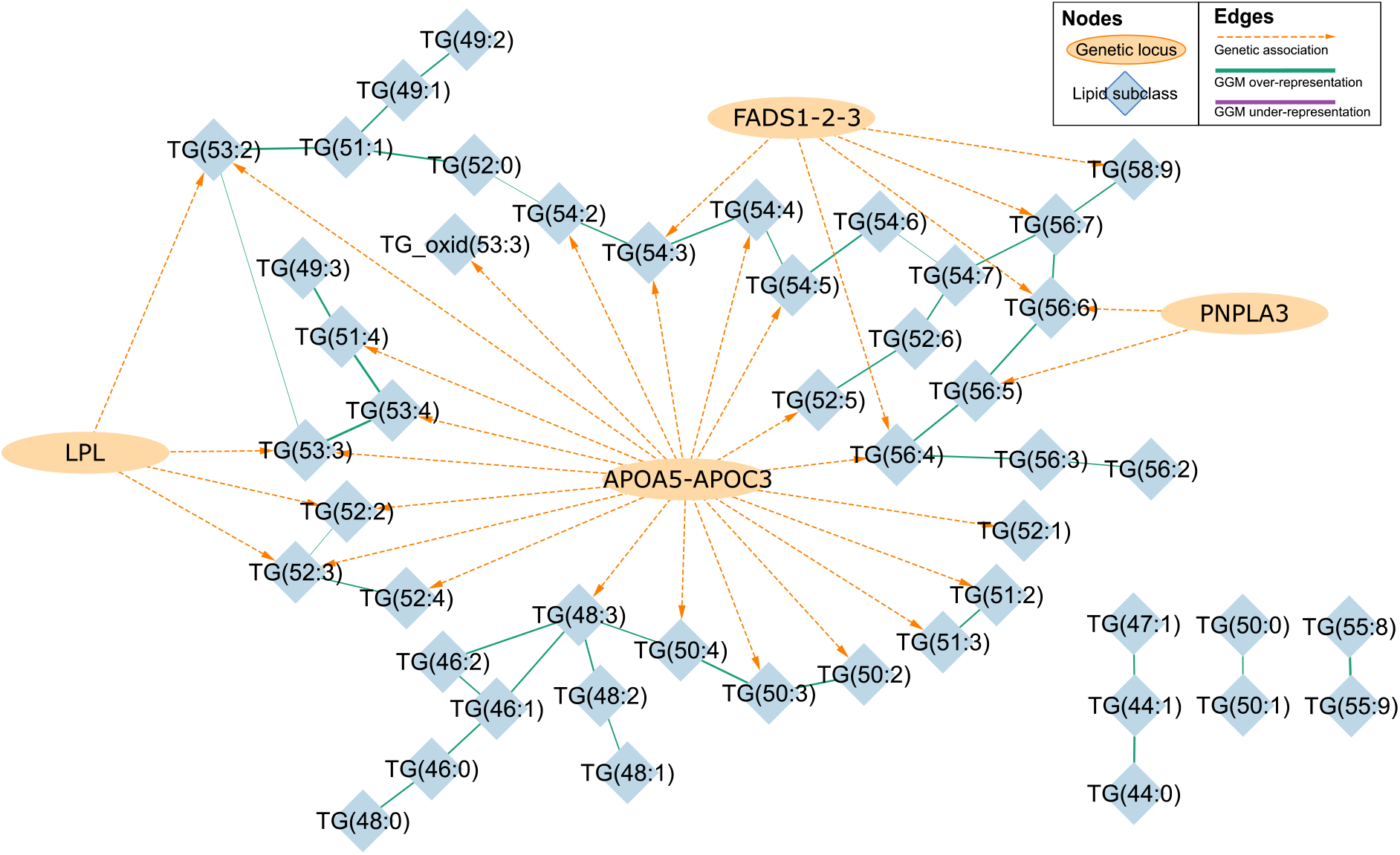
Combined network graph summarising genetic associations and a Gaussian graphical model (GGM) relating to levels of individual triglycerides in PROMIS. Nodes representing genetic loci are each labelled with the most likely “causal” gene at that locus according to our functional annotation (see Methods). In order for an edge to be drawn between a genetic locus and a triglyceride, there must have been a minimum of one variant at that locus significantly (*P* < 8.9 x 10^−10^) associated with at least one triglyceride. Edges between triglycerides indicate whether there was either a significant over- (green) or under- (purple) representation, with the magnitude indicated by the thickness of the edges.

## RESULTS

### Genetic architecture of the lipidome in South Asians and in the UK

We performed a genome-wide association study (GWAS) on the levels of 360 lipid metabolites using 6.7 million imputed autosomal variants in 5,662 hospital-based controls from PROMIS. We applied DIHRMS to quantify serum lipid metabolites across five broad lipid categories, i.e. fatty acyls and derivatives, glycerolipids, glycerophospholipids, sphingolipids, and sterol lipids [6]. We demonstrated the robustness of these lipid measurements in several ways, including validation of lipid signals against blanks, pooled samples, and internal standards, as we described previously [6]. Additionally, we replicated known associations of lipid metabolites with previously reported major lipid loci (Supplementary Table 15). After Bonferroni correction for multiple testing of variants and lipid metabolites (*P* < 8.929 × 10^−10^), we found 359 significant associations between 255 lipid metabolites and 24 genomic regions (Figure 1, Figure 2, Supplementary Figure 1, Supplementary Table 2). The majority of these lipid metabolites (67%; n = 171) were associated with variation at a single locus, while 26% of lipid metabolites were associated with two loci and 7% were associated with three or more loci (Supplementary Figures 2a and 3). To detect multiple independent associations at the same locus, we used stepwise conditional analysis, identifying 90 conditionally independent variants associated with lipid metabolites (Supplementary Table 3). 335 (93%) of the lipid QTLs had multiple conditionally significant associations (Supplementary Figure 2b).

Using the same DIHRMS platform, we also performed a GWAS on levels of 432 lipid metabolites using 87.7 million imputed autosomal variants in 13,814 British blood donors from INTERVAL. We identified significant associations with lipids at 38 independent loci (Figure 1, Supplementary Table 4). There was considerable consistency in the genomic regions identified in each study, with 18 (75%) of the significant genetic loci from PROMIS also found in INTERVAL (Figure 1). Six genetic loci were specific to lipid levels in the Pakastani population: *ANGPTL3, UGT8, PCTP, C19orf80, XBP1*, and *GAL3ST1*. There were also twenty genetic loci associated with lipids in the British population that were not significantly associated with lipids in the Pakistani population.

In PROMIS, the median proportion of variation in the lipidome explained by the genome-wide significant conditionally independent variants was 1.7% (interquartile range: 1.5-1.9%) (Supplementary Figure 2c), which is slightly less than that reported in metabolomics studies [16–19] but similar to the reported variation explained in previous lipidomics studies [20, 21]. There was a strong inverse relationship between effect size and minor allele frequency (MAF) (Supplementary Figure 2d), consistent with previous GWAS of quantitative traits [22, 23]. Approximately 70% of the analysed genetic variants in this analysis were common (MAF >5%) and 30% were low-frequency (MAF: 1-5%) with a median MAF of 8%. To help identify candidate causal genes through which genetic loci may influence lipid levels and thereby impact disease risk, we applied the ProGeM framework [11] (Supplementary Tables 7-13, Supplementary Figure 5). We identified a plausible or established link to biochemical function for 16 of the 24 loci (including *GCKR, LPL, FADS1-2-3*, and *APOA5-C3*), involving 34 unique genes. In cases where it was not possible to annotate SNPs using our systematic approach, we assigned them to their nearest protein-coding gene.

Previous studies have shown that the ratios of metabolites can strengthen association signals and lead to a better understanding of possible mechanisms [16]. Thus, in addition to the individual lipid metabolites, we selected twenty-six ratios of lipid metabolites that act through well-understood metabolic pathways. These included ratios associated with lipase activity, elongases, docosahexaenoic acid (DHA) levels, dairy fat intake, insulin production, glucose control, *de novo* lipogenesis, and cardiovascular disease risk (Supplementary Table 5). Genome-wide association analyses of these ratios in PROMIS resulted in the identification of four additional loci that were not detected in the GWAS of individual lipid metabolites (*MYCL1-MFSD2A, LPGAT1, LOC100507470*, and *HAPLN4-TM6SF1*) (Supplementary Table 6).

### Network of genetic and metabolic associations

To identify and visualise the connectivity between lipid subclasses, we generated a network of genetic and metabolic associations in PROMIS by summarising within each subclass the pairwise partial correlations between lipid metabolites and their genetic associations (Figure 3). This network diagram highlights that the number of connections between diglycerides and triglycerides was strongly over-represented in the Gaussian Graphical Model (GGM), indicating that there were more significant partial correlations between lipids from these subclasses than would be expected due to chance alone, whereas the number of connections between sphingomyelins and triglycerides was strongly under-represented in the GGM. In addition to being associated with variants from the *SPTLC3* and *FADS1-2-* 3 loci, we found that sphingomyelins were associated exclusively with four loci that were not associated with any other lipid subclasses: *GCKR, SGPP1, MLXIPL*, and *XBP1*.

Given the striking findings for triglycerides in the overall network diagram, we also generated a network in PROMIS for a subset of the triglyceride species showing the partial correlations of individual triglycerides and their detailed associations with genetic loci (Figure 4). This network diagram shows that variants in the *APOA5-C3* locus are associated with a wide range of triglycerides, consistent with previous associations of Apolipoprotein A-V (ApoA5) with plasma triglyceride levels. ApoA5 is a component of a number of lipoprotein fractions including HDL, VLDL, and chylomicrons, and it may regulate the catabolism of triglyceride-rich lipoprotein particles by *LPL* and/or play a role in the assembly of VLDL particles [24–28]. The network mainly shows links with triglycerides containing polyunsaturated fatty acids (PUFAs), suggesting that variants in the *APOA5-C3* locus mainly affect the catabolism of lipoproteins containing triglycerides derived from adipose tissues that are relatively enriched in more unsaturated fatty acids. In contrast, we did not see direct links of fully saturated triglycerides with the *APOA5-C3* locus, suggesting that genetic variation at this locus is not particularly involved in the assembly of VLDL particles in the liver as part of *de novo* lipogenesis (see Supplementary Figure 4). Fatty acid desaturase is key in the production of PUFAs; therefore, differences in *FADS1-2-3* activity are expected to be observed in triglycerides with a large number of double bonds and carbon atoms. Indeed, the GGM concords with established biochemistry since this locus is associated with triglycerides (TG) 56:6, 56:7, and 58:9 but is not associated with triglycerides with fewer double bonds or carbon atoms. In contrast, it is unclear why variants in the *PNPLA3* locus also have the strongest associations with triglycerides with a relatively larger number of carbon atoms and double bonds, namely TG(56:5) and TG(56:6) (see also Figure 5). One possible explanation is that significantly associated variants in the *PNPLA3* locus are changing the substrate specificity so that there is a shift in the relative amounts of triglycerides that are exported from the liver.

**Figure 5.**
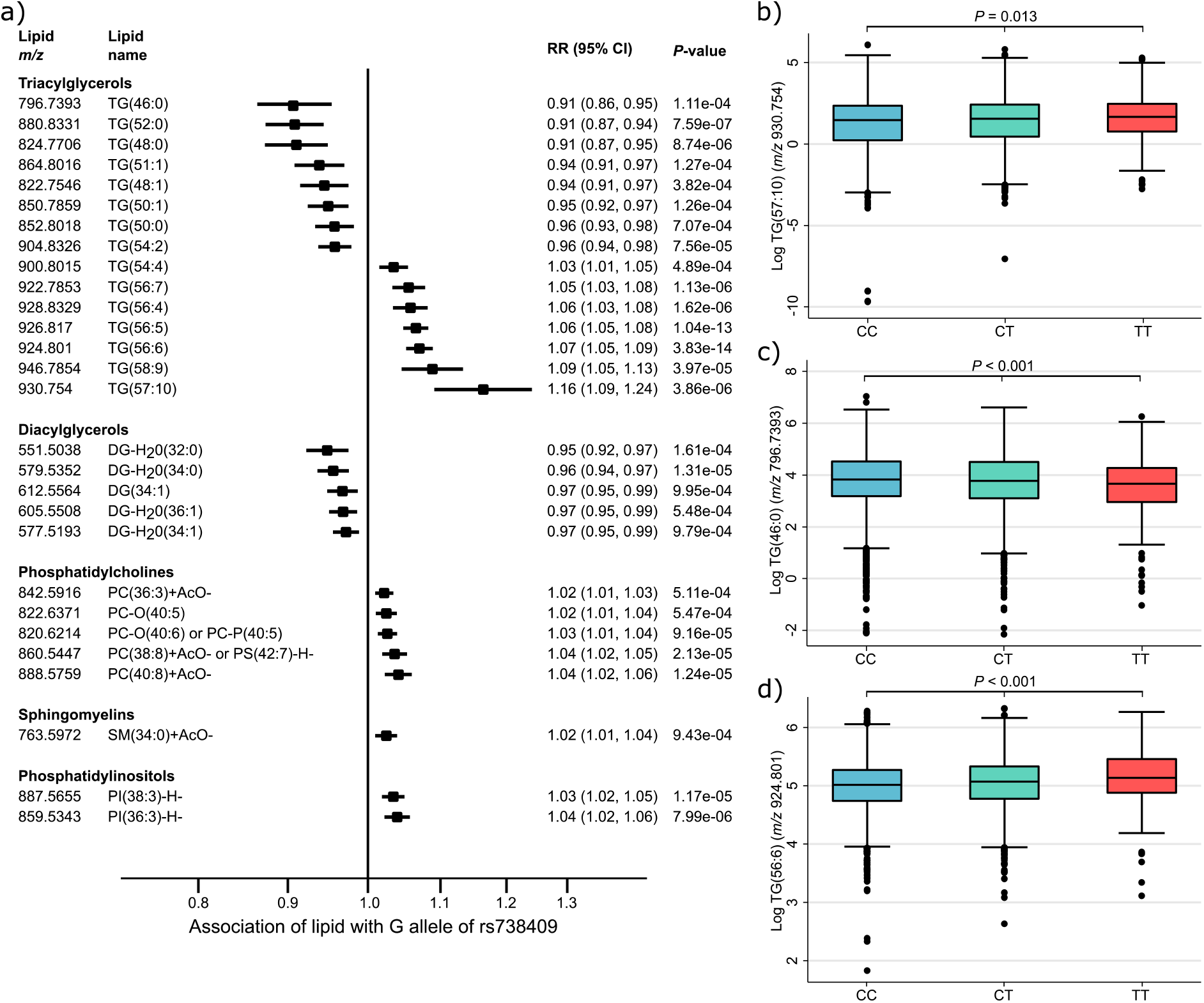
Association of lipids in PROMIS with *PNPLA3* and differences in levels of triglycerides by genotype. (a) Association of G allele of rs738409 in *PNPLA3* locus with levels of various lipids in PROMIS. The black lines denote 95% confidence intervals. Difference in levels of triglycerides in PROMIS by genotype: (b) TG(57:10) (*m/z* 930.754), (c) TG(46:0) (*m/z* 796.7393), and (d) TG(56:6) (*m/z* 924.801). *P*-values are for ANOVA test of difference in mean levels of triglycerides by genotype.

Additionally, the network diagram confirms that *LPL* is mainly active on MUFAs in triglyceride species. Variants in the *LPL* locus are significantly associated with TG(52:2), TG(52:3), TG(53:2), and TG(53:3), which have a high probability of containing one or more MUFAs within their fatty acid side chains. Figure 2 also shows that triglycerides and diglycerides are predominantly inversely associated with *LPL* variants, while triglycerides are positively associated with *PNPLA3* variants. Variants in the *LPL* locus are also positively associated with phosphocholines, sphingomyelins, and cholesterol esters, although the associations for the majority of the lipids in these subclasses did not reach genome-wide significance.

### New biological insights into lipid metabolism

Our analysis replicated known associations between lipids and genetic loci while also further extending what is known about these loci. We found significant associations of a wide range of lipids, including phosphatic acid (PA) 39:1, phosphatidylcholine (PC) 35:4, and phosphatidylethanolamines (PE) 36:4, 36:5, and 38:6, with variants in the *LIPC* locus (Supplementary Figure 6i); and significant associations of six specific sphingomyelins [SM(34:0), SM(40:0), SM(40:1), SM(40:2), SM(42:0)+AcO^-^, and SM(42:1)] and two phosphatidylcholines [PC-O(37:1) and PC-O(39:1)] with variants in the *APOE-C1-C2-C4* locus (Supplementary Figure 6b). We also identified significant associations of four further sphingomyelins [SM(31:1)-H^-^, SM(32:1), SM(32:1)+AcO^-^, and SM(39:1)] with variants in the *SGPP1* locus (Supplementary Figure 6s). Additionally, we found significant associations of nine ceramides [Cer(40:0)-H^-^, Cer(40:1)-H^-^, Cer(40:2)-H^-^, Cer(41:0)-H^-^, Cer(41:1)-H^-^, Cer(41:2)-H^-^, Cer(42:0)-H^-^, Cer(42:1)-H^-^, and Cer(42:2)-H^-^] with variants in the *SPTLC3* locus, which have not previously been reported in relation to this locus, as well as significant associations with three phosphatidylcholines and fifteen sphingomyelins (Supplementary Figure 6t).

We also discovered genetic associations with lipids at the patatin-like phospholipase domain containing protein 3 (*PNPLA3*) and membrane bound *O*-acyltransferase domain containing 7 (*MBOAT7*) loci that may have important biological and clinical implications. We found significant associations of two triglycerides—TG(56:6) (*m/z* 924.801) and TG(56:5) (*m/z* 926.817)—with rs12484809, an intronic variant in the *PNPLA3* locus (Supplementary Figure 6q). We also we found that the lead SNP in the *MBOAT7* locus, rs8736 (chr19:54677189), was associated with a wide range of phosphatic acids [e.g. PA(40:5) and PA(44:6)], phosphatidylcholines [e.g. PC(36:6) and PC(42:11)], phosphatidylethanolamines [e.g. PE(39:7)], and phosphoinositols [e.g. PI(34:1) and PI(36:1)] (Supplementary Figure 6k).

We undertook further investigation of a related nonsynonymous *PNPLA3* variant that is in moderate LD (*r*^2^ = 0.695), rs738409 (p.Ile148Met), to study the associations of lipids with *PNPLA3* in greater detail, including those that did not reach genome-wide significance. We focused on this variant rather than rs12484809 because I148M is already known to be associated with total triglycerides [29] and has been extensively characterised in previous genetic and functional analyses, and therefore is more likely to have potential clinical applications. As shown in Figure 5a, the *PNPLA3* I148M allele was associated with increased levels of lipids of higher carbon number and double-bond content, and consistently, with decreased levels of lipids of lower carbon number and double-bond content. There were also significant differences between the mean levels of the triglycerides TG(57:10), TG(46:0), and TG(56:6) between individuals stratified by *PNPLA3* I148M genotype (Figures 5b, 5c, and 5d).

## DISCUSSION

Based on a comprehensive analysis of genetic influences on 360 human blood lipids assayed in 5,662 individuals from Pakistan, we identified 359 significant associations between 255 lipids and 24 genetic loci. Additionally, in our analysis of 432 lipids in 13,814 British blood donors, we identified significant associations between 326 lipids and 38 independent loci. The majority of genetic regions associated with lipids in PROMIS were also found in INTERVAL; those that did not replicate may be due to the increased sample size in INTERVAL which gave a substantial boost in power. These findings suggest that genetically determined aspects of lipid metabolism are broadly similar in individuals of South Asian and European ancestry, and that DIHRMS can reliably capture differences in lipid levels across diverse populations.

There were six genetic loci specific to lipid levels in PROMIS: *ANGPTL3, UGT8, PCTP, C19orf80, XBP1*, and *GAL3ST1*. Angiopoietin-like 3 (*ANGPTL3*) is involved in regulation of lipid and glucose metabolism. SNPs in the *ANGPTL3* region have previously been shown to be associated with major lipids, including LDL-C and total cholesterol [30, 31]. In PROMIS, rs6657050, an intronic variant in the *ANGPTL3* locus, was significantly associated with PC(38:7)+AcO^-^ (*m/z* 862.5603) and PI(36:2)-H^-^ (*m/z* 861.5498) (Supplementary Figure 6a).

UDP glycosyltransferase 8 (*UGT8*) catalyses the transfer of galactose to ceramide, a key enzymatic step in the biosynthesis of galactocerebrosides, which are abundant sphingolipids of the myelin membrane of the central and peripheral nervous system. In PROMIS, rs28870381, an intergenic variant in *UGT8*, was associated with PG(32:1) (*m/z* 779.5078) (Supplementary Figure 6u).

Phosphatidylcholine transfer protein (*PCTP*) catalyses the transfer of phosphatidylcholines between membranes and is involved in lipid binding. Through regulation of plasma lipid concentrations it may also modulate the development of atherosclerosis [32]. In PROMIS, rs11079173, an intronic variant in the *PCTP* locus, was associated with PA(40:5)+AcO^-^ or PG(39:5)-H^-^ (*m/z* 809.5337) (Supplementary Figure 6n).

*C19orf80*, also known as angiopoietin-like 8 (*ANGPTL8*), is involved in the regulation of serum triglyceride levels, and is associated with major lipids including HDL-C and triglycerides [31]. In PROMIS, rs8101801, an intronic variant in the *C19orf80* locus, was significantly associated with PC(40:9)+AcO^-^ (*m/z* 886.5603) and PI(38:4)-H^-^ (*m/z* 885.5498) (Supplementary Figure 6d).

Galactose-3-*O*-sulfotransferase 1 (*GAL3ST1*) catalyses the sulfation of membrane glycolipids and the synthesis of galactosylceramide sulfate, a major lipid component of the myelin sheath. In PROMIS, rs2267161, a missense variant in the *GAL3ST1* locus, was associated with PG(32:1) (*m/z* 779.5078) (Supplementary Figure 6g).

X-box binding protein 1 (*XBP1*) functions as a transcription factor during endoplasmic reticulum stress by regulating the unfolded protein response. It is also a major regulator of the unfolded protein response in obesity-induced insulin resistance and T2D for the management of obesity and diabetes prevention. Recent studies have shown that compounds targeting the *XBP1* pathway are a potential approach for the treatment of metabolic diseases [33]. In addition, *XBP1* protein expression, which is induced in the liver by a high carbohydrate diet, is directly involved in fatty acid synthesis through *de novo* lipogenesis. Therefore, compounds that inhibit *XBP1* activation may also be useful for treatment of NAFLD [34]. In PROMIS, rs71661463, an intronic variant for which *XBP1* is the candidate causal gene, was associated with SM(37:1) (*m/z* 745.6216) (Supplementary Figure 6v). Recent research across many species has shown that *XBP1* is a transcription factor regulating hepatic lipogenesis. In mice, hepatic *XBP1* expression is regulated by proopiomelanocortin (POMC) during sensory food perception and coincides with changes in the lipid composition of the liver with increases in PCs and PEs [35]. Although previous studies have shown direct links between *XBP1* and overall lipid metabolism, this is the first time a genetic association has been reported between *XBP1* and lipid metabolites in humans, affecting sphingomyelins, PCs, and PEs (Supplementary Figure 6v).

Our findings for the *PNPLA3* and *MBOAT7* loci were also notable. *PNPLA3* is a multifunctional enzyme that encodes a triacylglycerol lipase, which mediates triacylglycerol hydrolysis in adipocytes and has acylglycerol *O*-acyltransferase activity. The relationship between rs738409, a nonsynonymous variant (p.Ile148Met) in the *PNPLA3* gene, and non-alcoholic fatty liver disease (NAFLD) has been well established [36]. This variant has been shown to impair triglyceride hydrolysis in the liver and secretion of triglyceride-rich very low density lipoproteins, leading to altered fatty acid composition of liver triglycerides, and is also associated with reduced risk of CHD [37] and increased risk of type 2 diabetes (T2D) [38]. This suggests that targeting hepatic pathways to reduce cardiovascular risk may be complex, despite the clustering of cardiovascular and hepatic diseases in people with metabolic syndrome. Our analysis offers granularity to the previously identified total triglyceride associations with *PNPLA3* by identifying two specific triglyceride species that may have a role in *PNPLA3* function.

*MBOAT7*, which contributes to the regulation of free arachidonic acid in the cell through the remodelling of phospholipids, was reported as being associated with the metabolite 1-arachidonoylglycerophosphoinositol in a previous mGWAS [16] [known as PI(36:4) in our study], but we found that the lead SNP in this locus, rs8736 (chr19:54677189), was also associated with a wide range of phosphatic acids, phosphatidylcholines, phosphatidylethanolamines, and phosphoinositols (Supplementary Figure 6k). Several studies have shown that *MBOAT7* (also known as lysophosphatidylinositol‐acyltransferase 1 [*LPIAT1*]) is responsible for the transfer of arachidonoyl‐CoA to lysophosphoinositides [39]. The creation of *MBOAT7*-deficient macrophages show a decreased level of PI(38:4) and an increase of PI(34:1) as well as PI(40:5) [40]. The T allele of rs8736, a 3’ UTR SNP, shows a similar shift in the phosphatidylinositol metabolism. Our work shows that this SNP is also strongly associated with PI(38:3), which is likely to be the dihomo-gamma linoleic acid (20:3n6)-containing phosphoinositol. None of the papers testing the substrate specificity of *MBOAT7* have included dihomo-gamma linoleic acid or PI(38:3) in their analysis. Thus, we provide novel evidence in humans that there is an association between *MBOAT7* activity and circulating phosphatidylinositols, a finding that requires further replication.

Our network diagram helped identify sphingomyelins that were associated exclusively with four loci that were not associated with any other lipid subclasses: *GCKR, SGPP1, MLXIPL*, and *XBP1*. Sphingomyelins have previously been shown to be associated with *SGPP1* [41], but the associations of sphingomyelins with these other three loci are reported here for the first time. *GCKR* has been shown to be associated with total cholesterol and triglycerides (see Figure 2), and has also been associated with the plasma phospholipid fraction fatty acids 16:0 and 16:1 [42, 43]; most lipids that we found to be associated with *GCKR* (Supplementary Figure 6g) are likely to contain these particular fatty acids. It has been suggested that the glucokinase receptor, encoded by *GCKR*, affects the production of malonyl-CoA, an important substrate for *de novo* lipogenesis [42]. To a similar extent there is a known relation between *MLXIPL* and carbohydrate and lipid metabolism. *MLXIPL* is a transcription factor affecting carbohydrate response element binding protein (CREBP) and therefore also plays a role in lipogenesis. Although both these genes have previously been linked to lipogenesis, we discovered that genetic variation at genes involved in the regulation of lipogenesis have been implicated in altering sphingomyelin concentrations.

The network diagram also helped recapitulate known biological relationships between lipids. As we established in our previous analysis [6], the number of significant partial correlations between lipids of different subclasses was significantly higher than would be expected due to chance alone. This analysis further showed that genes that were significantly associated with lipids of a particular subclass regulated all of the lipids within the subclass in a similar manner. Therefore, the total concentrations of a given lipid class associated with a genetic locus are less affected by the proportion of fatty acids present in those lipid species.

In summary, our analyses resulted in the following new insights in an understudied South Asian population: (1) we established that decreased levels of sphingomyelins are associated with genetically lower *LPL* activity; (2) we revealed a wide range of glycerophospholipids that are associated with variants in the *MBOAT7* locus; (3) we identified several new associations of phosphatic acids, phosphocholines, and phosphoethanolamines with variants in the *LIPC* region; (4) we found several novel associations of sphingomyelins and phosphocholines with variants in the *APOE-C1-C2-C4* cluster; (5) we discovered four new associations of sphingomyelins with variants in the *SGPP1* locus; and (6) we found several previously unreported associations of phosphocholines, sphingomyelins, and ceramides with variants in the *SPTLC3* locus. These findings can help further the identification of novel therapeutic targets for prevention and treatment.

Our investigation into the genetic influences of lipids has several strengths. First, the research involved participants from a population cohort in Pakistan, thereby enhancing scientific understanding of lipid associations in this understudied population, and we compared the findings with a typical Western population of British blood donors using the same lipid-profiling platform. Second, the analysis was based on a relatively large dataset of 5,662 participants from Pakistan and an even larger cohort of 13,814 individuals from the UK, thereby increasing statistical power to detect associations. Third, our mGWAS was performed in individuals free from established MI at baseline in PROMIS and healthy blood donors in INTERVAL, which reduces spurious associations due to the disease state or potential treatments. Finally, our newly developed open-profiling lipidomics platform was utilised to provide detailed lipid profiles, with a wider coverage of lipids than most other high-throughput profiling methods [6], which improved our ability to detect novel associations and our understanding of the detailed effects of known lipid loci at the level of individual lipid species.

Nevertheless, our study has several potential limitations. First, possible selection biases arise from the case-control design of PROMIS, although this was minimised by the recruitment of controls from patients, visitors of patients attending out-patient clinics, and unrelated visitors of cardiac patients. Second, serum samples in PROMIS were stored in freezers at −80 °C for between two to eight years before aliquots were taken for the lipidomics measurements, which we accounted for by adjusting the analyses by the number of years that the samples had been stored. Although residual confounding and deterioration of lipid profiles may still exist, such deterioration is unlikely to have been related to genotype. Third, a majority (76%) of PROMIS participants had not fasted prior to blood draw, and a small proportion of participants (7%) had reportedly fasted for an unknown duration. Recent food consumption may have had significant effects on lipid levels and influenced the results. Our analyses adjusted for fasting status although we lacked statistical power to stratify by fasting status. Fourth, PROMIS participants were recruited from multiple centres in urban Pakistan [6], but it is unclear whether the findings from this study would be generalizable to individuals living in rural villages and other parts of Pakistan, or in other countries in South Asia. However, the confirmatory analysis in INTERVAL, in which we identified significant associations with lipids for the majority of the genetic loci found in PROMIS, helps strengthen the argument that these findings are generalizable. Additionally, many of the lipids were associated with known genetic regions such as *APOA5-C3* and *FADS1-2-3*, which have already been shown to be associated with multiple lipids in other Western populations, further strengthening the validity of the findings from this analysis. Finally, although two-sample Mendelian randomization approaches to make causal inferences about the association of lipids with CHD risk factors and disease outcomes holds great promise in the lipidomics arena [44], extensive pleiotropy made it too difficult to disentangle the findings and we chose not to pursue this avenue. Therefore, although especially stringent procedures were followed, highly conservative cut-offs were used to determine statistical significance, and rigorous pre-analysis and post-analysis quality control steps were performed, there is still a possibility that some of the findings were false positives that arose due to artefacts rather than being true signals. Additional analyses in other populations using the DIHRMS lipidomics platform would be helpful to further replicate our findings. Moreover, the identified pathways and proposed molecular mechanisms require validation through functional analyses in model organisms and humans.

Further research will be able to leverage these lipidomics results in combination with whole-genome and whole-exome sequencing performed in PROMIS and INTERVAL to help understand the consequences of loss-of-function mutations identified in these participants [45].

## CONCLUSIONS

In conclusion, this article presents the results from a comprehensive analysis of genetic influences on human blood lipids in South Asians with a comparative analysis in the UK. Our findings strengthen and expand the knowledge base for understanding the genetic determinants of lipids and their association with cardiometabolic disease-related loci. These findings have important implications for the identification of novel therapeutic targets and advancement of mechanistic understanding of metabolic pathways that may lead to the onset of chronic diseases and lipid-related abnormalities.

## Supporting information

Additional file 3 - High-resolution version of Supplementary Figure 1

## Data Availability

The datasets used and/or analysed during the current study are available from the corresponding author on reasonable request.

## LIST OF ABBREVIATIONS

CHD: Coronary heart disease
CVD: Cardiovascular disease
DHA: Docosahexaenoic acid
DIHRMS: Direct infusion high resolution mass spectrometry
FDR: False discovery rate
GGM: Gaussian Graphical Model
HWE: Hardy-Weinberg Equilibrium
MAF: Minor allele frequency
MI: Myocardial infarction
*m/z*: Mass-charge ratio
NAFLD: Non-alcoholic fatty liver disease
PROMIS: Pakistan Risk of Myocardial Infarction Study
PUFA: Polyunsaturated fatty acid
SD: Standard deviation
SNP: Single nucleotide polymorphism
T2D: Type 2 diabetes
QC: Quality Control
QTL: Quantitative trait loci

## DECLARATIONS

### Ethics approval and consent to participate

#### PROMIS

The institutional review board at the Center for Non-Communicable Diseases in Karachi, Pakistan approved the study (IRB: 00007048, IORG0005843, FWAS00014490) and all participants gave informed consent, including for use of samples in genetic, biochemical, and other analyses.

#### INTERVAL

The National Research Ethics Service approved this study (11/EE/0538) and all participants gave electronic informed consent.

### Consent for publication

Not applicable.

### Funding

#### PROMIS

Fieldwork, genotyping, and standard clinical chemistry assays in PROMIS were principally supported by grants awarded to the University of Cambridge from the British Heart Foundation (SP/09/002; RG/13/13/30194), the UK Medical Research Council (G0800270; MR/L003120/1), the Wellcome Trust, the EU Framework 6–funded Bloodomics Integrated Project, Pfizer, Novartis, and Merck.

#### INTERVAL

Participants in the INTERVAL randomised controlled trial were recruited with the active collaboration of NHS Blood and Transplant England (http://www.nhsbt.nhs.uk), which has supported field work and other elements of the trial. DNA extraction and genotyping was co-funded by the National Institute for Health Research (NIHR), the NIHR BioResource (http://bioresource.nihr.ac.uk), and the NIHR [Cambridge Biomedical Research Centre at the Cambridge University Hospitals NHS Foundation Trust] [*]. The academic coordinating centre for INTERVAL was supported by core funding from: NIHR Blood and Transplant Research Unit in Donor Health and Genomics (NIHR BTRU-2014-10024), UK Medical Research Council (MR/L003120/1), British Heart Foundation (SP/09/002, RG/13/13/30194; RG/18/13/33946) and the NIHR [Cambridge Biomedical Research Centre at the Cambridge University Hospitals NHS Foundation Trust] [*]. A complete list of the investigators and contributors to the INTERVAL trial is provided in reference [46]. The academic coordinating centre would like to thank blood donor staff and blood donors for participating in the INTERVAL trial. [*] The views expressed are those of the authors and not necessarily those of the NHS, the NIHR, or the Department of Health and Social Care.

This work was supported by Health Data Research UK, which is funded by the UK Medical Research Council, Engineering and Physical Sciences Research Council, Economic and Social Research Council, Department of Health and Social Care (England), Chief Scientist Office of the Scottish Government Health and Social Care Directorates, Health and Social Care Research and Development Division (Welsh Government), Public Health Agency (Northern Ireland), British Heart Foundation, and Wellcome.

J.L.G. and A.K. are funded by the UK Medical Research Council under the Lipid Dynamics and Regulation supplementary grant (MC_PC_13030) and Lipid Programming and Signalling program grant (MC_UP_A090_1006) and Cambridge Lipidomics Biomarker Research Initiative (G0800783). D.S.P. and D.St. are funded by the Wellcome Trust (105602/Z/14/Z).

### Authors’ contributions

E.L.H., J.D., D.Sa., J.L.G., and A.K. conceived and designed the study. J.D. and D.Sa. are principal investigators of PROMIS. A.M.W., J.L.G., and A.K. jointly supervised the research. A.K. generated the lipidomics data. E.L.H. and A.K. processed the lipidomics data. E.L.H. performed the bioinformatics and statistical analyses. E.B.F., D.St., D.S.P., D.Z., R.M.Y.O., A.S.B., A.M.W., J.L.G., and A.K. contributed important intellectual content to the study and manuscript. E.L.H., A.S.B., A.M.W., J.L.G., and A.K. were involved in drafting the manuscript. All authors read and approved the final manuscript.

## Acknowledgements

The authors would like to thank Michael Inouye for his helpful comments on an earlier version of the manuscript.

## SUPPLEMENTARY INFORMATION

**Additional file 1**. Supplementary Methods; Supplementary Figures 1-6; References for Supplementary Material

**Additional file 2**. Supplementary Tables 1-15

**Additional file 3**. Supplementary Figure 1 (high resolution)

