## Additional file 1 - Supplementary Methods & Figures for "Genome-wide analysis of blood lipid metabolites in over 5,000 South Asians reveals biological insights at cardiometabolic disease loci"

**SUPPLEMENTARY MATERIAL**

**Supplementary Methods**

Lipid profiling, genotyping, and genetic analyses in INTERVAL

Further details of candidate gene annotation in PROMIS and INTERVAL

**Supplementary Tables**

Supplementary Table 1. Number of significant variants and loci associated with each lipid subclass

Supplementary Table 2. Summary of significant associations between lipid metabolites and genetic variants from univariate genome-wide association study in PROMIS

Supplementary Table 3. Summary of significant associations between lipid metabolites and genetic variants from conditional analyses of univariate genome-wide association study results in PROMIS

Supplementary Table 4. Summary of significant associations between lipid metabolites and genetic variants from univariate genome-wide association study in INTERVAL

Supplementary Table 5. Summary and classification of analysed ratios in PROMIS

Supplementary Table 6. Summary of significant associations between ratios of lipid metabolites and genetic variants from univariate genome-wide association study in PROMIS

Supplementary Table 7. Prediction of causal genes in PROMIS based on integration of information from bottom-up and top-down SNP annotation approaches

Supplementary Table 8. Annotation of genome-wide significant associations in PROMIS from the level of the variant (bottom-up)

Supplementary Table 9. Annotation of genome-wide significant associations in PROMIS by proximal (±500-Kb from lead variant) gene function (top-down)

Supplementary Table 10. Lead variants in PROMIS residing in exonic sequence

Supplementary Table 11. Lead variants in PROMIS in high linkage disequilibrium ($r^2 \geq 0.8$) with ≥ 1 non-synonymous SNP

Supplementary Table 12. Lead SNP *cis*-eQTLs in PROMIS in lipid-relevant human tissues

Supplementary Table 13. Enrichment analysis of cell-type specific enhancer overlap with our set of 90 lead SNPs from conditional analyses in PROMIS using HaploReg v4.1

Supplementary Table 14. Prediction of causal genes in INTERVAL based on integration of information from bottom-up and top-down SNP annotation approaches

Supplementary Table 15. Summary of associations between lipid metabolites in PROMIS and 175 major lipid loci

**Supplementary Figures**

Supplementary Figure 1. Extended heat map showing associations of significant loci from conditional analyses with all lipid metabolites, major lipids, and lipid-related diseases/disorders

Supplementary Figure 2. Genetic architecture of serum lipid levels

Supplementary Figure 3. Number of lipids associated with each variant

Supplementary Figure 4. Increased *de novo* lipogenesis in lipodystrophy and NAFLD patients

Supplementary Figure 5. Flow diagram outlining strategy for mediating gene prioritisation

Supplementary Figure 6. Association of lipids with significantly associated loci from conditional analyses

#### SUPPLEMENTARY METHODS

##### Lipid profiling, genotyping, and genetic analyses in INTERVAL

INTERVAL is a cohort study of nearly 50,000 healthy blood donors in the UK, the details of which have been described previously [1]. In this study we analysed data from 13,814 participants with genetic and lipid-profiling data.

We performed DIHRMS on INTERVAL participants using the same protocol that we used for PROMIS, as described previously [2]. We obtained data on 432 lipids, 253 in positive ionisation mode and 179 in negative ionisation mode, which make up 19 lipid subclasses (Supplementary Table 1).

DNA was extracted from buffy coat at LGC Genomics (UK) using a Kleargene method and samples of sufficient concentration and purity were aliquoted for shipment to Affymetrix for genotyping [3]. Duplicate samples and samples that were not of European ancestry were excluded. Additionally, SNPs were excluded if (1) the variant had fewer than 10 called minor allele homozygotes, (2) the cluster plot contained at least one sample with an intensity at least twice as far from the origin as the next most extreme sample, (3) the outlying sample(s) had an extreme polar angle ($<15^\circ$ or $>75^\circ$) in the direction of the minor allele, (4) call rate < 99% per batch and < 75% overall, (5) MAF < 0.05, (6) HWE $P < 1 \times 10^{-6}$, or (7) $r^2 \geq 0.2$ between pairs of variants [3]. The dataset was phased using SHAPEIT3 (in chunks of 5,000 variants with an overlap of 250 variants between chunks) and subsequently imputed using the 1000 Genomes Phase 3-UK10K imputation panel, resulting in 87,696,910 imputed variants in the dataset [3]. In total, 13,814 individuals from INTERVAL had overlapping information on lipidomics data and imputed SNPs.

Linear regression was performed using BOLT-LMM v2.2 [4] to determine the association of each lipid with each SNP. Residuals were calculated from the null model for each lipid with adjustment for plate, age, sex, centre, appointment month, appointment hour, processing time in hours, and the first three genetic principal components. The threshold for genome-wide significance level was set to $P < 4.464 \times 10^{-10}$ ($5 \times 10^{-8}$ / 112), as 112 principal components explained >95% of the variance in lipid levels.

**Further details of candidate gene annotation in PROMIS and INTERVAL**

To prioritise candidate genes underlying the associations with lipid metabolites in PROMIS and INTERVAL, we applied the ProGeM framework, which combines information from complementary "bottom-up" and "top-down" approaches to assess the credibility of potential candidate genes [5]. The process described below is for PROMIS, though we also used the same approach for INTERVAL.

For the bottom-up approach, we annotated SNPs based on their putative effects on proximal gene function if any of the following conditions were met: (1) the SNP resided within an exonic sequence of a gene (Supplementary Table 9); (2) the SNP resided within a splice-site (±2-bp from an intron-exon boundary); (3) the SNP was in high linkage disequilibrium (LD) ($r^2 \geq 0.8$) with a non-synonymous SNP (Supplementary Table 10); and/or (4) the SNP was a *cis*-eQTL for a local gene (Supplementary Table 11). To identify any exonic and splice site SNPs within our SNP list, we ran the Variant Effect Predictor (VEP) (http://www.ensembl.org/common/Tools/VEP?db=core) on the list of variants with the "pick" option (which outputs one block of annotation per variant) and used Ensembl transcripts as the reference for determining consequences. SNPs in high LD with our list of associated SNPs were identified within our imputed dataset and run through VEP to select only non-synonymous SNPs. *Cis*-eQTLs within our list of associated SNPs were identified using eQTL data provided by the Genotype-Tissue Expression (GTEx) project (http://www.gtexportal.org/home/datasets), keeping only significant SNP-gene associations (filename: "GTEx_Analysis_v7_eQTL.tar.gz"). We only annotated SNPs if they were significant eQTLs in at least one of the following tissues we deemed most relevant for lipid-related phenotypes: subcutaneous adipose tissue, visceral adipose tissue, liver, and/or whole blood.

In the top-down approach, for each of our associated SNPs, we first identified all proximal genes located ≤ 500-Kb upstream or downstream using the ANNOVAR tool (http://annovar.openbioinformatics.org/en/latest/). We then identified all genes previously associated with a lipid-related biological process or function from the following databases: (1) LIPID MAPS Proteome Database (LMPD) (http://www.lipidmaps.org/data/proteome/LMPD.php); (2) Gene Ontology (GO) (http://geneontology.org/); (3) Online Mendelian Inheritance in Man catalogue (OMIM); (4) Mouse Genome Informatics (MGI) database (http://www.informatics.jax.org/); (5) Kyoto Encyclopedia of Genes and Genomes (KEGG) (http://www.genome.jp/kegg/); and/or (6) Ingenuity Pathway Analysis (IPA) (http://www.ingenuity.com/products/ipa).

LMPD is an object-relational database of lipid-associated genes and proteins across multiple species including human, mouse, and fruit fly; we simply extracted all human genes (1,116 genes in total) from this database (accessed 16-Mar-2016). For GO and OMIM, we first identified all terms or Mendelian diseases containing one or more lipid-related keyword(s) using HumanMine (http://www.humanmine.org/), then we extracted all human genes associated with one or more of these terms (accessed 01-Apr-2016 and 07-Apr-2016). Similarly, for MGI we extracted all mouse genes using MouseMine (http://www.mousemine.org/mousemine/begin.do) (accessed 31-Mar-2016) that were associated with the following manually-selected lipid-related terms and their children: (1) abnormal lipid homeostasis (MP:0002118); (2) abnormal lipoprotein level (MP:0010329); (3) abnormal lipid metabolism (MP:0013245); and (4) adipose tissue phenotype (MP:0005375). From the KEGG database we extracted all lipid compounds (with "C" number IDs) with biological roles in order to identify all genes associated with reactions (with "R" number IDs) involved in lipid biology using MitoMiner (http://mitominer.mrc-mbu.cam.ac.uk/release-3.1/begin.do) (accessed 31-Mar-2016). Finally, from IPA we downloaded the interaction networks for all fourteen of the lipid subclasses in order to extract all genes in a compound-specific manner (accessed 13-Apr-2016).

Once we had obtained lists of lipid-related genes from the aforementioned databases, we then searched for overlap with our list of proximal (≤ 500-Kb) genes based on HUGO Gene Nomenclature Committee (HGNC) symbols, thereby annotating SNPs with proximal genes where there was evidence that at least one might be involved in lipid-related biology. For each lead SNP we first recorded whether there was any compound-specific evidence from IPA for a SNP-gene assignment whereby both the SNP (from this study) and the gene (from IPA) were associated with the same lipid subclass. Then from the five remaining (compound non-specific) databases we categorised overlapping genes as either (1) recurrent candidates, in that they were highlighted in at least two different databases, or (2) single candidates. Further, we assigned the recurrent candidates a score out of five for prioritisation purposes, with one point awarded for each database highlighting them as being lipid-related.

### SUPPLEMENTARY FIGURES

**Supplementary Figure 1.** Extended heat map showing associations of significant loci from conditional analyses with all lipid metabolites, major lipids, and lipid-related diseases/disorders

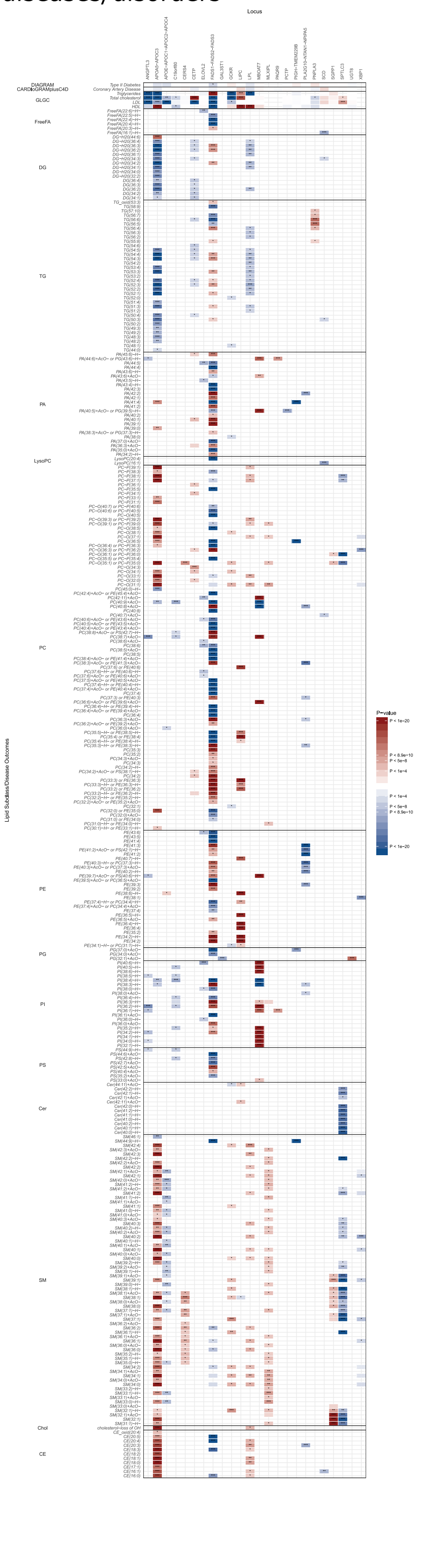

**Caption for Supplementary Figure 1 (previous page):**
The effect estimates of the associations between significant variants with all lipid metabolites are plotted as a heat map. Results are shown for the association of all lipid metabolites (rows) with the most strongly associated genetic variant within each locus (columns). The associations with major lipids from the GLGC (total cholesterol, HDL-C, LDL-C, and triglycerides), DIAGRAM Consortium (type 2 diabetes), and CARDIoGRAMplusC4D Consortium (coronary artery disease) are also shown. The magnitude and direction of the effect estimates (standardised per 1-SD) are indicated by a colour scale, with blue indicating a negative association and red indicating a positive association with respect to the SNP effect on the trait. Asterisks indicates the degree of significance of the *P*-values of association. * = $P < 1 \times 10^{-4}$; ** = $P < 5 \times 10^{-8}$; *** = $P < 8.9 \times 10^{-10}$. **Note:** A high-resolution version of this figure is available as Additional file 3.

**Supplementary Figure 2.** Genetic architecture of serum lipid levels

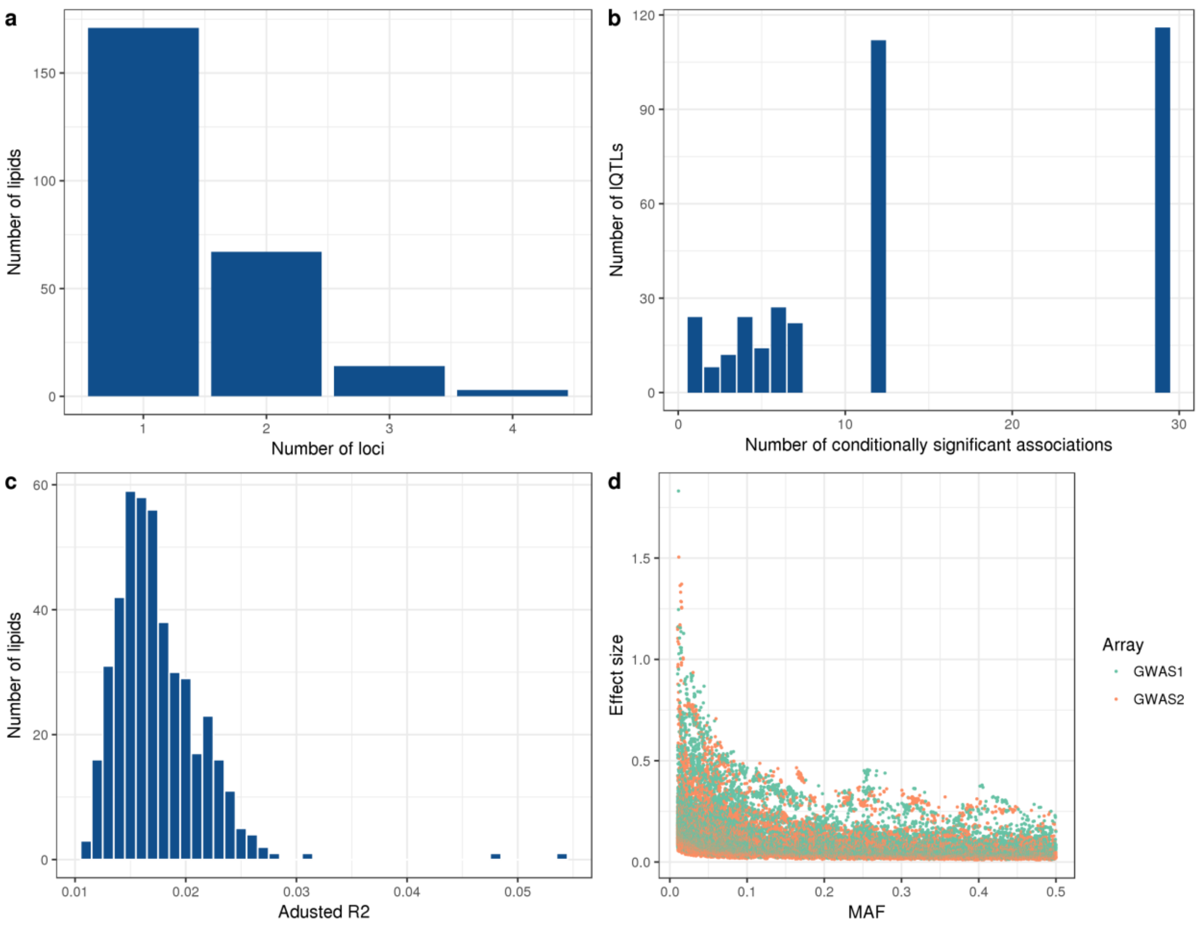


n = 5,662 participants. (a) Number of significantly associated loci per lipid. (b) Number of conditionally significant associations within each lipid QTL. (c) Histogram of variance explained by conditionally independent variants. (d) Effect size versus MAF.

**Supplementary Figure 3.** Number of lipids associated with each variant

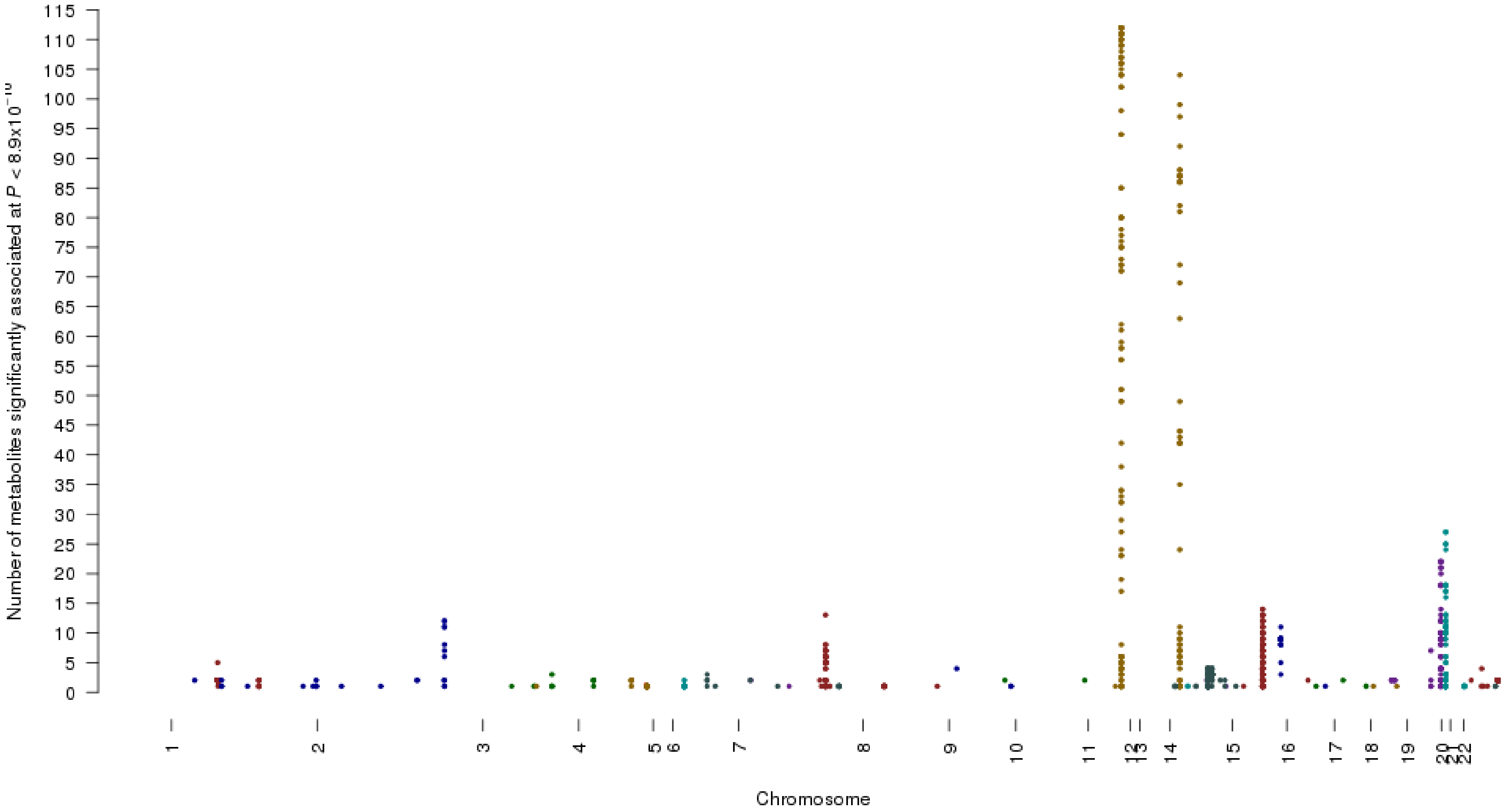


Results are shown for the number of lipids associated with each variant at genome-wide significance ($P < 8.9 \times 10^{-10}$).

**Supplementary Figure 4.** Increased *de novo* lipogenesis in lipodystrophy and NAFLD patients

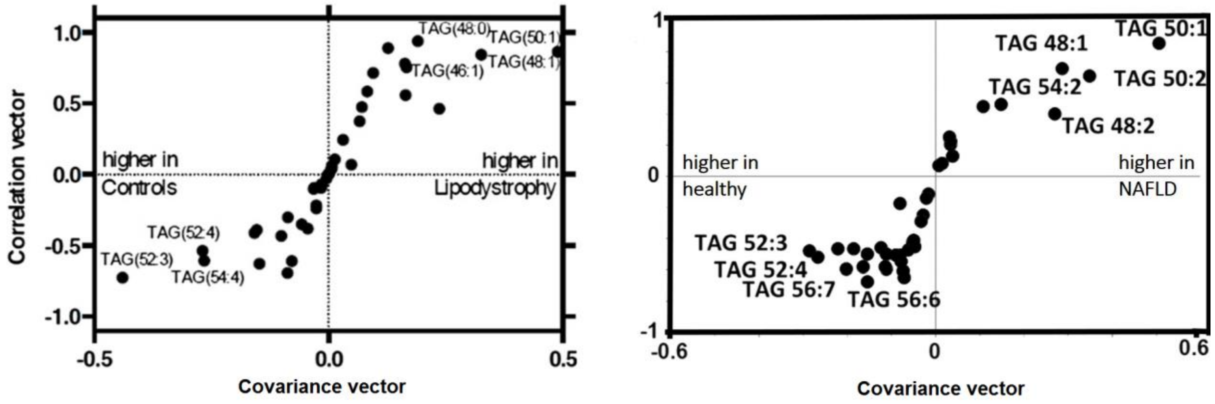


Increased *de novo* lipogenesis in lipodystrophy patients (left panel based on Eiden et al 2015) [6] and NAFLD patients (right panel based on Sanders et al 2018) [7] both show an increase in TAG(48:1) and TG(50:1) originating from the liver, leading to lower levels of TG(52:4) and other triglycerides associated with *APOA5-C3* variants.

**Supplementary Figure 5.** Flow diagram outlining strategy for mediating gene prioritisation

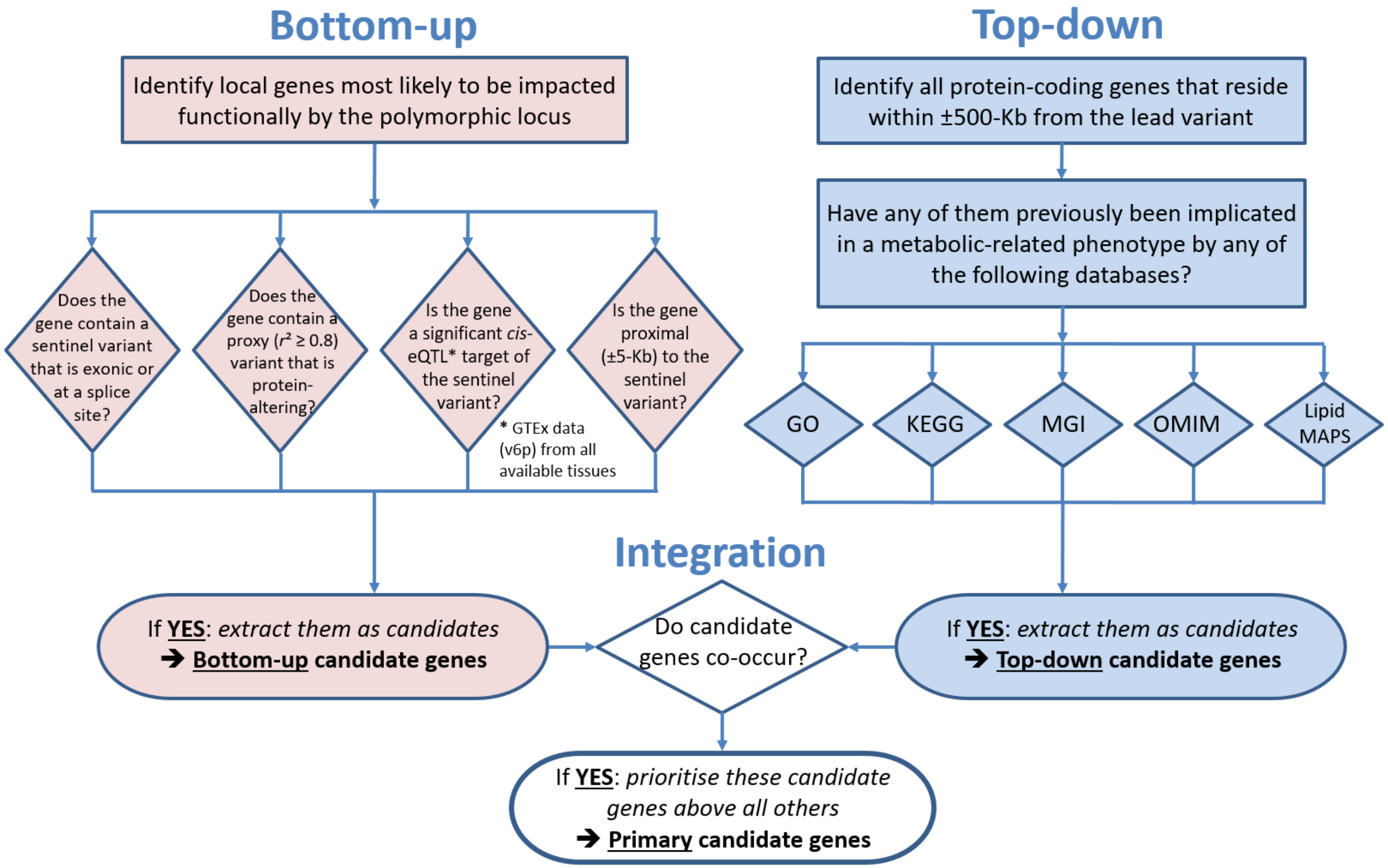


The flow diagram shows how the "bottom-up" and "top-down" approaches were used and then integrated to identify probable causal genes for each significantly associated variant. A proxy is defined as those variants with $r^2 \geq 0.8$ with the lead (EUR population, 1000 Genomes). **Abbreviations: eQTL** = Expression Quantitative Trait Locus; **GO** = Gene Ontology; **GTEx** = Genotype-Tissue Expression; **KEGG** = Kyoto Encyclopedia of Genes and Genomes; **Lipid MAPS** = Lipid Metabolites and Pathways Strategy; **MGI** = Mouse Genome Informatics; **OMIM** = Online Inheritance in Man. Adapted from Stacey et al [5].

**Supplementary Figure 6.** Association of lipids with significantly associated loci from conditional analyses

#### (a) *ANGPTL3*

| Lipid name | Lipid *m/z* | Beta (95% CI) | *P*-value |
|---|---|---|---|
| SM(36:1)+AcO- | 789.6128 | 0.02 (0.01, 0.03) | 1.51e-04* |
| PC-O(33:1) | 732.5904 | 0.02 (0.01, 0.03) | 3.28e-05* |
| SM(36:1) | 731.6062 | 0.02 (0.01, 0.03) | 1.13e-04* |
| PC(40:9)+AcO- | 886.5603 | -0.03 (-0.04, -0.02) | 2.04e-06* |
| PI(38:4)-H- | 885.5498 | -0.03 (-0.04, -0.02) | 1.21e-06* |
| PI(40:5)-H- | 911.5655 | -0.03 (-0.04, -0.01) | 8.40e-05* |
| PS(44:9)-H- | 884.5448 | -0.03 (-0.05, -0.02) | 1.84e-05* |
| PI(34:1)-H- | 835.5341 | -0.03 (-0.05, -0.02) | 7.31e-05* |
| PI(38:5)-H- | 883.5343 | -0.03 (-0.05, -0.02) | 6.14e-06* |
| PC(38:8)+AcO- or PS(42:7)-H- | 860.5447 | -0.03 (-0.05, -0.02) | 4.37e-07* |
| PC(36:6)+AcO- or PE(39:6)+AcO- | 836.5446 | -0.04 (-0.05, -0.02) | 8.04e-05* |
| PI(34:2)-H- | 833.5186 | -0.04 (-0.05, -0.02) | 9.38e-08* |
| PI(36:3)-H- | 859.5343 | -0.04 (-0.05, -0.02) | 2.46e-07* |
| PE(39:7)+AcO- or PS(40:6)-H- | 834.529 | -0.04 (-0.05, -0.02) | 6.31e-08* |
| PI(36:1)-H- | 863.5654 | -0.04 (-0.06, -0.03) | 2.04e-07* |
| PA(44:6)+AcO- or PG(43:6)-H- | 863.5806 | -0.04 (-0.06, -0.03) | 1.88e-07* |
| PI(36:2)-H- | 861.5498 | -0.04 (-0.06, -0.03) | 1.63e-13*** |
| PC(38:7)+AcO- | 862.5603 | -0.05 (-0.06, -0.03) | 2.28e-13*** |
| PI(34:0)-H- | 837.5498 | -0.05 (-0.07, -0.02) | 4.58e-05* |
| PI(35:2)-H- | 847.5343 | -0.05 (-0.07, -0.03) | 5.58e-06* |

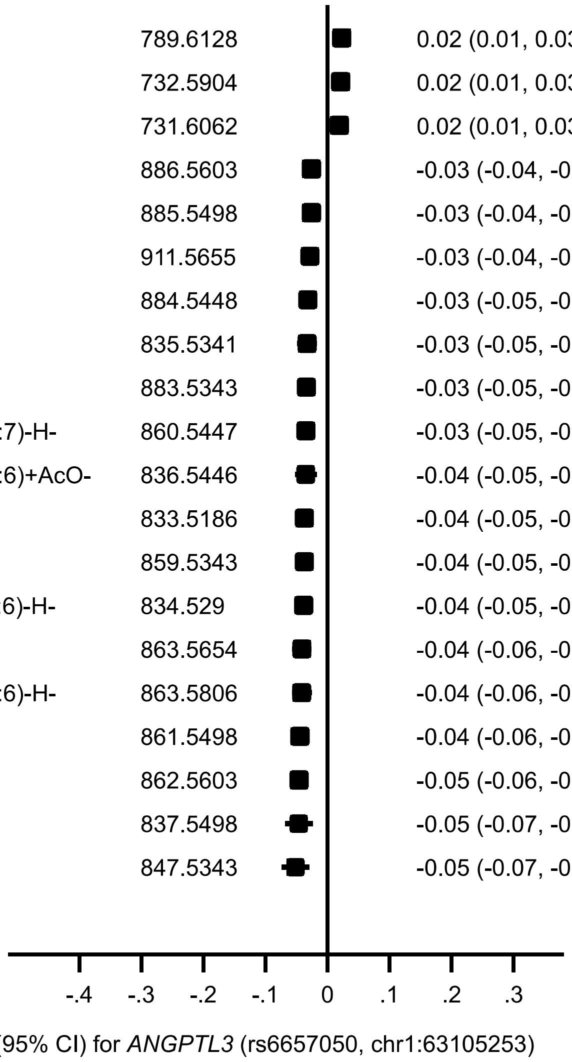

**(b) *APOA5-APOC3***

| Lipid name | Lipid *m/z* | Beta (95% CI) | *P*-value |
|---|---|---|---|
| CE(18:3) | 664.6026 | 0.08 (0.07, 0.10) | 4.28e-23*** |
| SM(42:2) | 813.6844 | 0.08 (0.06, 0.09) | 2.99e-23*** |
| PC-P(37:1) | 786.6373 | 0.07 (0.06, 0.09) | 4.06e-27*** |
| SM(40:2) | 785.6529 | 0.07 (0.06, 0.08) | 2.44e-27*** |
| PC-O(33:1) | 732.5904 | 0.07 (0.05, 0.08) | 3.31e-24*** |
| SM(36:1) | 731.6062 | 0.07 (0.05, 0.08) | 3.92e-26*** |
| PC-O(31:1) | 704.5591 | 0.07 (0.05, 0.08) | 1.06e-27*** |
| SM(34:0) | 705.5906 | 0.07 (0.05, 0.08) | 1.08e-26*** |
| PC-O(35:1) or PC-P(35:0) | 760.6219 | 0.07 (0.05, 0.08) | 9.66e-24*** |
| PC-O(37:1) | 788.653 | 0.07 (0.05, 0.08) | 1.13e-25*** |
| SM(38:1) | 759.6372 | 0.07 (0.05, 0.08) | 1.56e-24*** |
| SM(34:1) | 703.5747 | 0.07 (0.05, 0.08) | 2.85e-27*** |
| CE(18:0) | 670.6496 | 0.06 (0.05, 0.08) | 1.53e-24*** |
| SM(40:1) | 787.6688 | 0.06 (0.05, 0.08) | 1.35e-25*** |
| SM(40:0) | 789.6845 | 0.06 (0.05, 0.08) | 6.35e-25*** |
| CE(18:1) | 668.6339 | 0.06 (0.05, 0.07) | 9.11e-24*** |
| cholesterol-loss of OH | 369.3514 | 0.06 (0.05, 0.07) | 6.74e-25*** |
| DG-H20(36:2) | 603.5352 | -0.09 (-0.10, -0.07) | 2.06e-24*** |
| TG(52:3) | 874.7859 | -0.09 (-0.11, -0.07) | 4.17e-25*** |
| TG(54:3) | 902.8175 | -0.10 (-0.12, -0.08) | 3.18e-23*** |

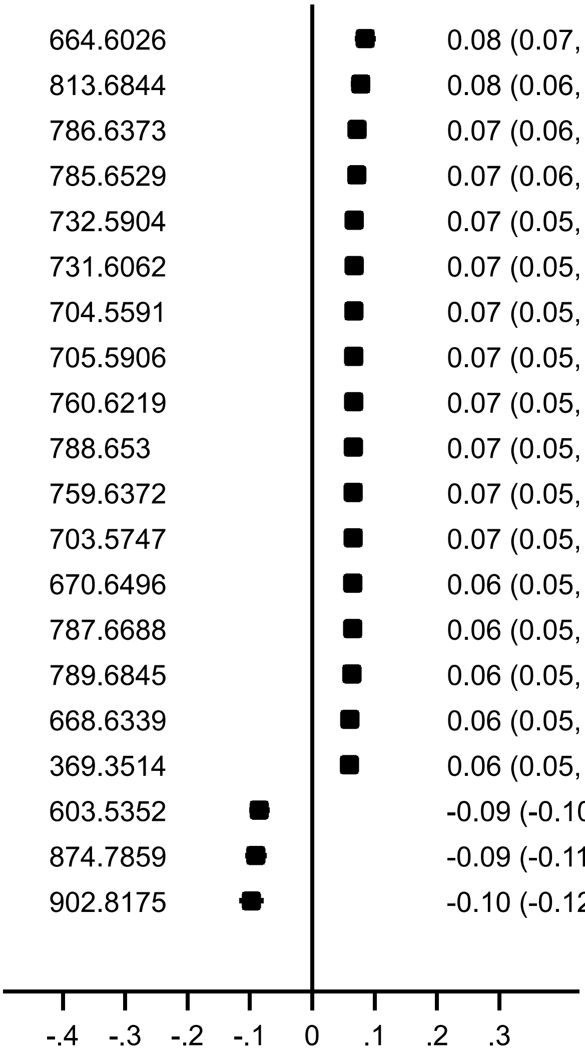


Beta (95% CI) for *APOA5-APOC3* (rs2072560, chr11:116661826)

**(c) *APOE-APOC1-APOC2-APOC4***

| Lipid name | Lipid *m/z* | Beta (95% CI) | *P*-value |
|---|---|---|---|
| SM(38:1)+AcO- | 817.6441 | -0.04 (-0.06, -0.03) | 1.05e-06* |
| SM(34:0)+AcO- | 763.5972 | -0.05 (-0.06, -0.03) | 8.72e-07* |
| SM(34:1)+AcO- | 761.5815 | -0.05 (-0.06, -0.03) | 2.42e-07* |
| SM(37:1)-H- | 743.6075 | -0.05 (-0.06, -0.03) | 3.14e-07* |
| SM(33:1)-H- | 687.5448 | -0.05 (-0.06, -0.03) | 4.64e-08** |
| SM(39:2)-H- | 769.6231 | -0.05 (-0.06, -0.03) | 1.05e-07* |
| SM(40:2)+AcO- | 843.6598 | -0.05 (-0.07, -0.03) | 5.19e-08* |
| SM(40:0)+AcO- | 847.6911 | -0.05 (-0.07, -0.03) | 1.74e-07* |
| SM(33:0)-H- | 689.5604 | -0.05 (-0.07, -0.03) | 4.68e-08** |
| SM(39:1)-H- | 771.6386 | -0.05 (-0.07, -0.03) | 2.47e-08** |
| SM(40:1)+AcO- | 845.6754 | -0.05 (-0.07, -0.03) | 1.77e-08** |
| SM(40:2)-H- | 783.6387 | -0.05 (-0.07, -0.03) | 8.38e-08* |
| SM(42:2)+AcO- | 871.6911 | -0.05 (-0.07, -0.03) | 4.94e-07* |
| SM(41:2)-H- | 797.6543 | -0.05 (-0.07, -0.03) | 2.00e-07* |
| SM(41:2)+AcO- | 857.6754 | -0.05 (-0.07, -0.04) | 5.12e-08* |
| SM(42:1)+AcO- | 873.7067 | -0.06 (-0.08, -0.04) | 5.20e-09** |
| SM(42:0)+AcO- | 875.7224 | -0.06 (-0.08, -0.04) | 8.24e-10*** |
| SM(39:0)-H- | 773.6543 | -0.06 (-0.08, -0.04) | 6.39e-09** |
| SM(41:1)-H- | 799.67 | -0.06 (-0.08, -0.04) | 1.20e-09** |
| SM(41:0)-H- | 801.6857 | -0.07 (-0.09, -0.04) | 1.71e-07* |

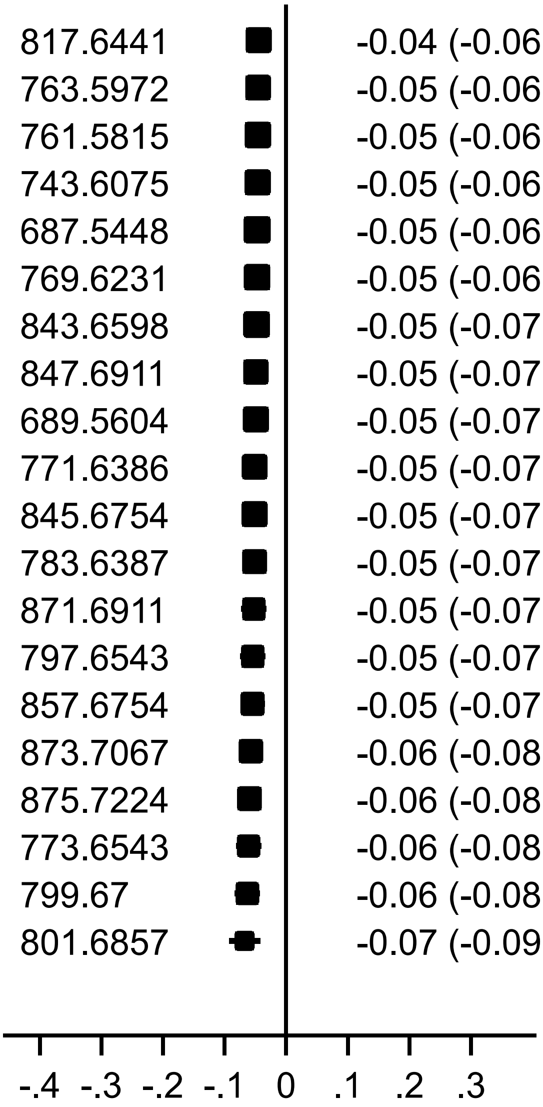


Beta (95% CI) for *APOE-APOC1-APOC2-APOC4* (rs75627662, chr19:45413576)

**(d) *C19orf80***

| Lipid name | Lipid *m/z* | Beta (95% CI) | *P*-value |
|---|---|---|---|
| SM(42:2)+AcO- | 871.6911 | 0.04 (0.02, 0.06) | 1.41e-05* |
| SM(42:2) | 813.6844 | 0.04 (0.02, 0.05) | 1.25e-05* |
| PC-P(39:1) | 814.6688 | 0.04 (0.02, 0.05) | 2.16e-05* |
| PC(38:8)+AcO- or PS(42:7)-H- | 860.5447 | -0.04 (-0.06, -0.02) | 4.34e-05* |
| PI(34:2)-H- | 833.5186 | -0.04 (-0.06, -0.02) | 3.76e-05* |
| PE(39:7)+AcO- or PS(40:6)-H- | 834.529 | -0.04 (-0.06, -0.02) | 3.83e-05* |
| PI(36:2)-H- | 861.5498 | -0.04 (-0.06, -0.03) | 5.26e-07* |
| PC(38:7)+AcO- | 862.5603 | -0.04 (-0.06, -0.03) | 4.94e-07* |
| PI(36:4)-H- | 857.5186 | -0.05 (-0.07, -0.03) | 1.70e-06* |
| PI(38:4)-H- | 885.5498 | -0.05 (-0.06, -0.03) | 3.56e-10*** |
| PC(40:8)+AcO- | 888.5759 | -0.05 (-0.07, -0.03) | 1.27e-05* |
| PC(40:9)+AcO- | 886.5603 | -0.05 (-0.06, -0.03) | 3.38e-10*** |
| PS(42:8)-H- | 858.5291 | -0.05 (-0.07, -0.03) | 6.80e-07* |
| PI(38:5)-H- | 883.5343 | -0.05 (-0.07, -0.03) | 4.23e-06* |
| TG(51:3) | 860.7703 | -0.05 (-0.07, -0.03) | 2.52e-05* |
| PS(44:9)-H- | 884.5448 | -0.05 (-0.07, -0.03) | 1.97e-06* |
| PI(40:5)-H- | 911.5655 | -0.05 (-0.07, -0.03) | 3.46e-07* |
| TG(53:4) | 886.7859 | -0.06 (-0.08, -0.03) | 3.55e-06* |
| PI(35:2)-H- | 847.5343 | -0.08 (-0.11, -0.05) | 5.88e-07* |
| TG(51:4) | 858.7544 | -0.08 (-0.12, -0.05) | 6.56e-06* |

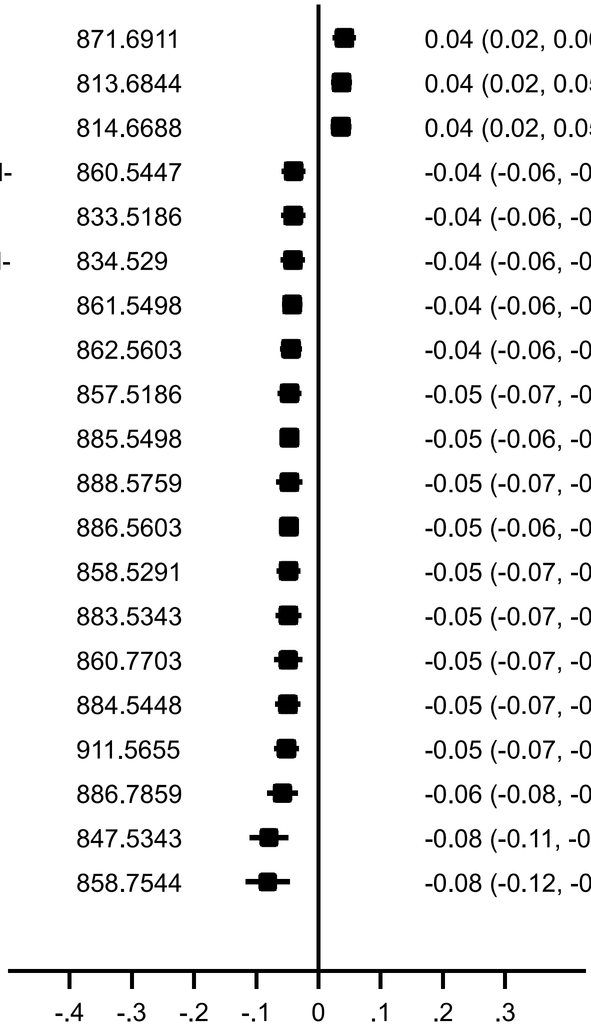


Beta (95% CI) for *C19orf80* (rs8101801, chr19:11335477)

**(c) *CERS4***

| Lipid name | Lipid *m/z* | Beta (95% CI) | *P*-value |
|---|---|---|---|
| SM(37:1)+AcO- | 803.6286 | 0.04 (0.03, 0.06) | 3.74e-08** |
| SM(38:0)+AcO- | 819.6598 | 0.04 (0.03, 0.05) | 1.74e-08** |
| SM(38:1) | 759.6372 | 0.04 (0.03, 0.05) | 1.65e-12*** |
| SM(37:1)-H- | 743.6075 | 0.04 (0.02, 0.05) | 6.76e-09** |
| SM(38:0) | 761.6532 | 0.04 (0.02, 0.05) | 2.43e-07* |
| PC-O(35:1) or PC-P(35:0) | 760.6219 | 0.03 (0.02, 0.04) | 8.53e-11*** |
| SM(38:1)+AcO- | 817.6441 | 0.03 (0.02, 0.05) | 5.44e-08* |
| SM(37:1) | 745.6216 | 0.03 (0.02, 0.05) | 1.41e-07* |
| SM(36:1)-H- | 729.5917 | 0.03 (0.02, 0.05) | 1.16e-05* |
| SM(35:2)-H- | 713.5605 | 0.03 (0.02, 0.05) | 5.28e-07* |
| SM(36:2) | 729.5906 | 0.03 (0.02, 0.04) | 2.41e-08** |
| SM(35:0)-H- | 717.5918 | 0.03 (0.02, 0.04) | 6.89e-06* |
| SM(35:1)-H- | 715.5761 | 0.03 (0.02, 0.04) | 8.75e-07* |
| SM(36:2)+AcO- | 787.5972 | 0.03 (0.02, 0.04) | 8.55e-06* |
| PC-O(33:1) | 732.5904 | 0.03 (0.02, 0.04) | 1.01e-07* |
| SM(36:1) | 731.6062 | 0.03 (0.02, 0.04) | 4.88e-08** |
| SM(36:1)+AcO- | 789.6128 | 0.03 (0.02, 0.04) | 1.33e-05* |
| PC-P(33:1) | 730.5747 | 0.03 (0.01, 0.04) | 4.11e-04* |
| SM(36:0) | 733.6219 | 0.02 (0.01, 0.03) | 7.01e-06* |
| SM(36:0)+AcO- | 791.6285 | 0.02 (0.01, 0.03) | 5.86e-05* |

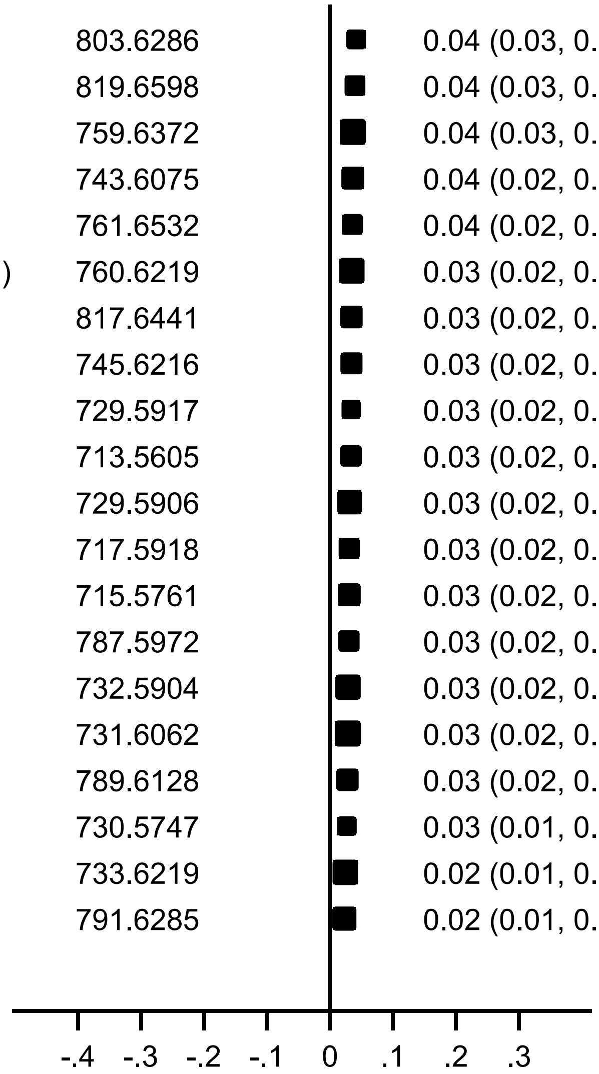


Beta (95% CI) for *CERS4* (rs11666866, chr19:8285607)

**(d) *CETP***

| Lipid name | Lipid *m/z* | Beta (95% CI) | *P*-value |
|---|---|---|---|
| PC-O(34:3) | 742.5748 | 0.04 (0.03, 0.05) | 4.08e-10*** |
| PC-P(36:1) | 772.6219 | 0.03 (0.02, 0.05) | 5.39e-06* |
| PC-P(34:1) | 744.5904 | 0.03 (0.02, 0.04) | 1.65e-06* |
| PC-O(34:1) | 746.6061 | 0.03 (0.02, 0.04) | 2.27e-06* |
| PA(40:1) | 759.5903 | 0.02 (0.01, 0.02) | 1.14e-06* |
| PC(34:2) | 758.57 | 0.02 (0.01, 0.02) | 2.22e-06* |
| DG-H20(36:2) | 603.5352 | -0.03 (-0.04, -0.02) | 7.96e-06* |
| DG(36:2) | 638.5723 | -0.03 (-0.05, -0.02) | 2.69e-06* |
| TG(56:6) | 924.801 | -0.03 (-0.05, -0.02) | 9.69e-06* |
| DG-H20(36:3) | 601.5195 | -0.03 (-0.05, -0.02) | 8.59e-06* |
| TG(54:4) | 900.8015 | -0.04 (-0.05, -0.02) | 1.04e-05* |
| DG(34:2) | 610.541 | -0.04 (-0.06, -0.02) | 5.53e-07* |
| TG(56:7) | 922.7853 | -0.04 (-0.06, -0.02) | 5.41e-06* |
| DG(34:1) | 612.5564 | -0.04 (-0.06, -0.03) | 5.17e-07* |
| TG(54:5) | 898.7856 | -0.04 (-0.06, -0.03) | 1.83e-06* |
| TG(52:5) | 870.7544 | -0.04 (-0.06, -0.03) | 7.25e-06* |
| SM(46:1) | 871.7627 | -0.05 (-0.06, -0.03) | 6.44e-06* |
| TG(54:6) | 896.7698 | -0.05 (-0.07, -0.03) | 1.45e-06* |
| DG(36:4) | 634.5409 | -0.05 (-0.07, -0.03) | 1.16e-06* |
| TG(54:7) | 894.7541 | -0.06 (-0.09, -0.04) | 5.02e-06* |

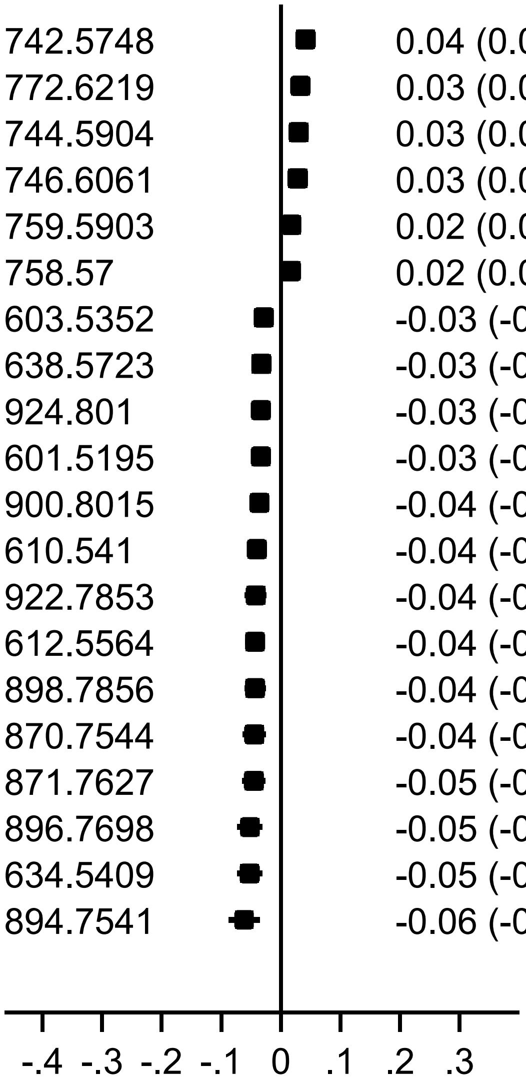


Beta (95% CI) for *CETP* (rs711752, chr16:56996211)

#### (e) *ELOVL2*

| Lipid name | Lipid *m/z* | Beta (95% CI) | *P*-value |
|---|---|---|---|
| CE(20:5) | 688.6026 | 0.04 (0.02, 0.06) | 1.47e-04* |
| PC(38:5)+AcO- | 866.5916 | 0.03 (0.01, 0.04) | 1.78e-05* |
| PC(38:5) | 808.5851 | 0.02 (0.01, 0.02) | 2.62e-04* |
| PG(36:2)-H- | 773.5337 | -0.02 (-0.04, -0.01) | 3.97e-04* |
| PC(37:6) or PE(40:6) | 792.5537 | -0.03 (-0.04, -0.01) | 2.21e-04* |
| PC(38:6) | 806.5694 | -0.03 (-0.04, -0.02) | 2.86e-09** |
| PI(38:6)-H- | 881.5186 | -0.03 (-0.05, -0.02) | 1.12e-04* |
| PA(44:5) | 807.5903 | -0.03 (-0.04, -0.02) | 3.57e-09** |
| PI(38:0)-H- | 893.6124 | -0.03 (-0.05, -0.02) | 1.25e-05* |
| PC(38:6)+AcO- | 864.5759 | -0.03 (-0.05, -0.02) | 2.92e-06* |
| PI(36:0)-H- | 865.5811 | -0.03 (-0.05, -0.02) | 1.51e-06* |
| PC(40:6)+AcO- or PE(43:6)+AcO- | 892.6072 | -0.03 (-0.05, -0.02) | 1.05e-05* |
| PE(43:6) | 834.6006 | -0.03 (-0.05, -0.02) | 8.50e-08* |
| PC(37:6)-H- or PE(40:6)-H- | 790.5391 | -0.04 (-0.05, -0.02) | 2.33e-07* |
| PE(38:6)-H- | 762.5079 | -0.04 (-0.05, -0.02) | 7.83e-05* |
| PC(37:6)+AcO- or PE(40:6)+AcO- | 850.5602 | -0.04 (-0.05, -0.02) | 6.57e-05* |
| PA(43:5)-H- | 791.5596 | -0.04 (-0.05, -0.02) | 9.10e-08* |
| FreeFA(22:6)-H- | 327.233 | -0.04 (-0.06, -0.03) | 1.68e-08** |
| PC(42:11)+AcO- | 910.5603 | -0.05 (-0.06, -0.03) | 8.02e-09** |
| PI(40:6)-H- | 909.5498 | -0.05 (-0.06, -0.03) | 1.43e-10*** |

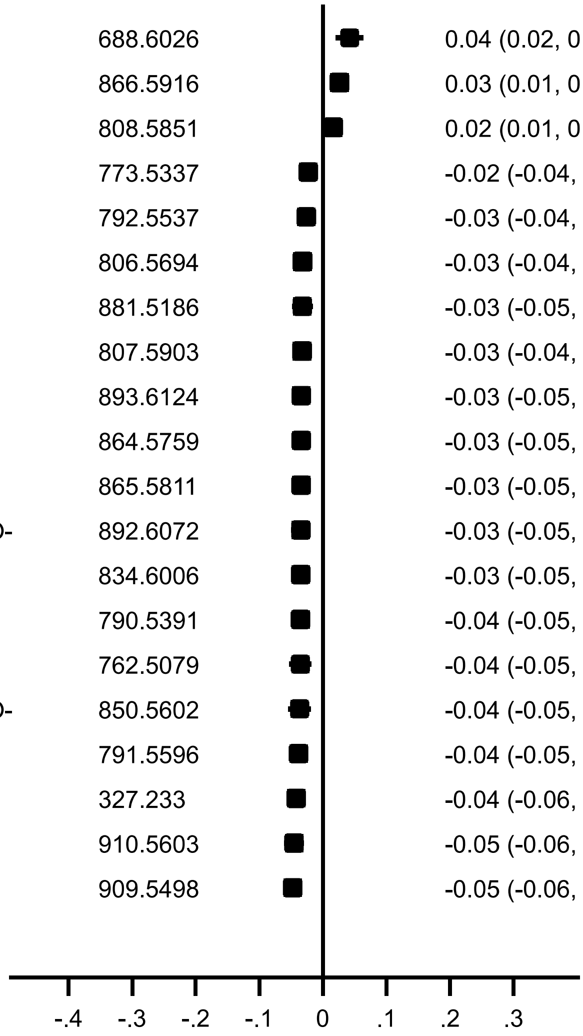


Beta (95% CI) for *ELOVL2* (rs6920155, chr6:11047956)

#### (f) *FADS1-FADS2-FADS3*

| Lipid name | Lipid *m/z* | Beta (95% CI) | *P*-value |
|---|---|---|---|
| PC(40:8)+AcO- | 888.5759 | 0.19 (0.17, 0.21) | 2.94e-84*** |
| PI(38:3)-H- | 887.5655 | 0.14 (0.12, 0.16) | 9.18e-68*** |
| PE(39:3) | 784.5855 | 0.07 (0.06, 0.08) | 9.96e-62*** |
| PA(42:2) | 785.6059 | 0.06 (0.05, 0.07) | 1.87e-46*** |
| PC(38:5) | 808.5851 | -0.08 (-0.09, -0.07) | 4.12e-48*** |
| PA(44:4) | 809.6059 | -0.09 (-0.10, -0.07) | 2.02e-51*** |
| PE(43:5) | 836.6163 | -0.11 (-0.12, -0.09) | 2.14e-47*** |
| PC(38:5)+AcO- | 866.5916 | -0.11 (-0.13, -0.10) | 8.95e-49*** |
| PC(35:4)-H- or PE(38:4)-H- | 766.5391 | -0.12 (-0.13, -0.10) | 1.17e-68*** |
| PC(36:4) | 782.5699 | -0.13 (-0.14, -0.12) | 4.8e-130*** |
| PC-O(36:5) | 766.5745 | -0.13 (-0.14, -0.11) | 4.38e-77*** |
| PA(42:3) | 783.5903 | -0.13 (-0.14, -0.12) | 4.7e-134*** |
| PC(37:4)-H- or PE(40:4)-H- | 794.5704 | -0.13 (-0.15, -0.11) | 7.38e-62*** |
| PA(41:4) | 767.559 | -0.13 (-0.14, -0.12) | 7.53e-81*** |
| PC(36:4)+AcO- or PE(39:4)+AcO- | 840.5759 | -0.13 (-0.15, -0.12) | 3.66e-79*** |
| PC(38:4)+AcO- or PE(41:4)+AcO- | 868.6072 | -0.14 (-0.16, -0.13) | 4.82e-74*** |
| PE(41:4) | 810.6012 | -0.15 (-0.16, -0.13) | 7.2e-115*** |
| SM(44:9)-H- | 825.5918 | -0.15 (-0.17, -0.13) | 1.06e-74*** |
| PC(37:4) | 796.5856 | -0.15 (-0.17, -0.13) | 3.30e-56*** |
| CE(20:4) | 690.6183 | -0.20 (-0.21, -0.18) | 1.2e-112*** |

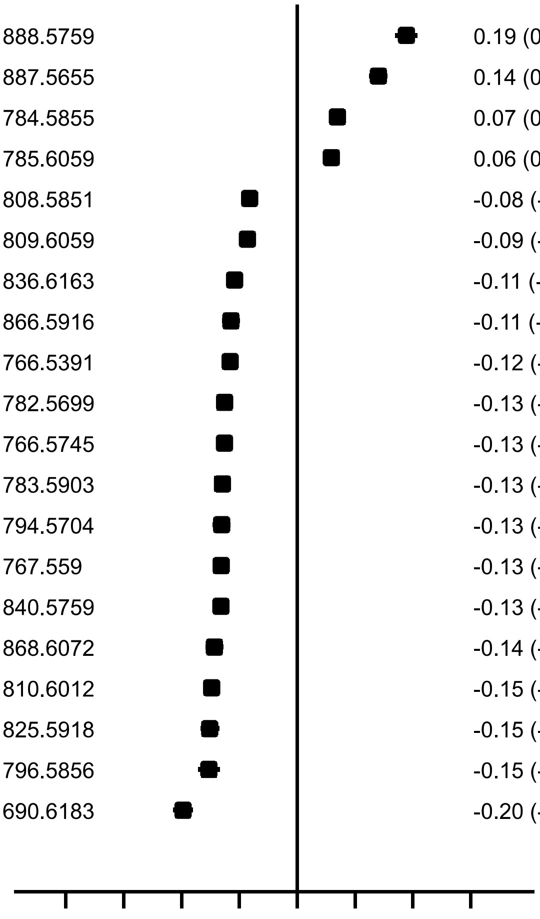


Beta (95% CI) for *FADS1-FADS2-FADS3* (rs174546, chr11:61569830)

#### (g) *GAL3ST1*

| Lipid name | Lipid *m/z* | Beta (95% CI) | *P*-value |
|---|---|---|---|
| TG(46:0) | 796.7393 | 0.06 (0.01, 0.11) | 1.24e-02 |
| DG-H20(30:1) | 521.4569 | 0.05 (0.01, 0.09) | 2.01e-02 |
| TG(46:2) | 792.7077 | 0.05 (0.01, 0.08) | 1.59e-02 |
| DG(34:2) | 610.541 | 0.03 (0.01, 0.05) | 1.57e-03 |
| DG(34:1) | 612.5564 | 0.03 (0.01, 0.05) | 5.77e-03 |
| TG(50:2) | 848.77 | 0.03 (0.01, 0.05) | 1.50e-02 |
| TG(50:3) | 846.7546 | 0.02 (0.00, 0.04) | 1.34e-02 |
| DG(36:2) | 638.5723 | 0.02 (0.01, 0.04) | 9.90e-03 |
| DG-H20(34:2) | 575.5039 | 0.02 (0.00, 0.04) | 1.37e-02 |
| PC(36:4) | 782.5699 | -0.01 (-0.02, -0.00) | 1.29e-02 |
| PA(42:3) | 783.5903 | -0.01 (-0.02, -0.00) | 1.32e-02 |
| SM(38:1) | 759.6372 | -0.01 (-0.03, -0.00) | 9.46e-03 |
| PE(41:4) | 810.6012 | -0.02 (-0.03, -0.00) | 7.18e-03 |
| PC(37:3) or PE(40:3) | 798.6012 | -0.02 (-0.03, -0.00) | 1.74e-02 |
| PE(38:1) | 774.6009 | -0.02 (-0.03, -0.00) | 1.36e-02 |
| PC(42:4)+AcO- or PE(45:4)+AcO- | 924.6698 | -0.02 (-0.04, -0.00) | 1.64e-02 |
| PC(37:4) | 796.5856 | -0.03 (-0.04, -0.01) | 4.34e-03 |
| SM(38:0) | 761.6532 | -0.03 (-0.04, -0.01) | 4.86e-04* |
| PS(40:1)+AcO- | 904.6284 | -0.07 (-0.11, -0.04) | 6.76e-05* |
| PG(32:1)+AcO- | 779.5078 | -0.12 (-0.16, -0.08) | 4.86e-10*** |

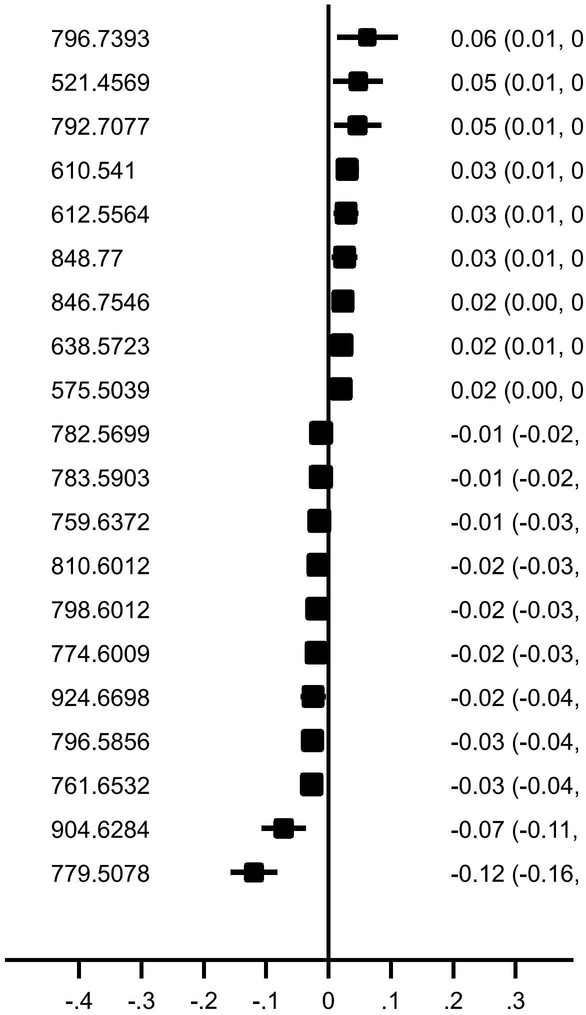


Beta (95% CI) for *GAL3ST1* (rs2267161, chr22:30953295)

**(h) *GCKR***

| Lipid name | Lipid *m/z* | Beta (95% CI) | *P*-value |
|---|---|---|---|
| SM(42:2)-H- | 811.67 | 0.05 (0.03, 0.07) | 4.09e-07* |
| SM(37:1) | 745.6216 | 0.05 (0.03, 0.06) | 5.47e-11*** |
| SM(36:1)-H- | 729.5917 | 0.05 (0.03, 0.06) | 4.99e-08** |
| SM(32:1)-H- | 673.5291 | 0.04 (0.03, 0.06) | 2.20e-10*** |
| SM(38:1)-H- | 757.623 | 0.04 (0.03, 0.06) | 5.10e-07* |
| PC-P(36:1) | 772.6219 | 0.04 (0.02, 0.06) | 5.32e-07* |
| PC-P(34:1) | 744.5904 | 0.04 (0.02, 0.05) | 1.27e-07* |
| SM(39:1) | 773.6531 | 0.04 (0.02, 0.05) | 4.66e-07* |
| PC-O(34:3) | 742.5748 | 0.04 (0.02, 0.05) | 8.22e-07* |
| PC-O(36:3) or PC-P(36:2) | 770.6063 | 0.04 (0.02, 0.05) | 1.17e-07* |
| PC-P(38:3) | 796.6219 | 0.04 (0.02, 0.05) | 6.35e-07* |
| PC-P(38:1) | 800.6529 | 0.03 (0.02, 0.04) | 4.32e-07* |
| SM(32:1) | 675.5434 | 0.03 (0.02, 0.04) | 9.71e-08* |
| SM(31:1)-H- | 659.5135 | 0.03 (0.02, 0.04) | 5.05e-07* |
| CE(18:2) | 666.6183 | 0.03 (0.02, 0.04) | 6.16e-07* |
| PC-O(35:1) or PC-P(35:0) | 760.6219 | 0.03 (0.02, 0.04) | 9.50e-07* |
| PC-O(31:1) | 704.5591 | 0.03 (0.02, 0.04) | 1.21e-07* |
| SM(34:0) | 705.5906 | 0.03 (0.02, 0.04) | 1.47e-07* |
| SM(34:1) | 703.5747 | 0.03 (0.02, 0.04) | 1.00e-07* |
| SM(34:2) | 701.5593 | 0.03 (0.02, 0.04) | 4.77e-07* |

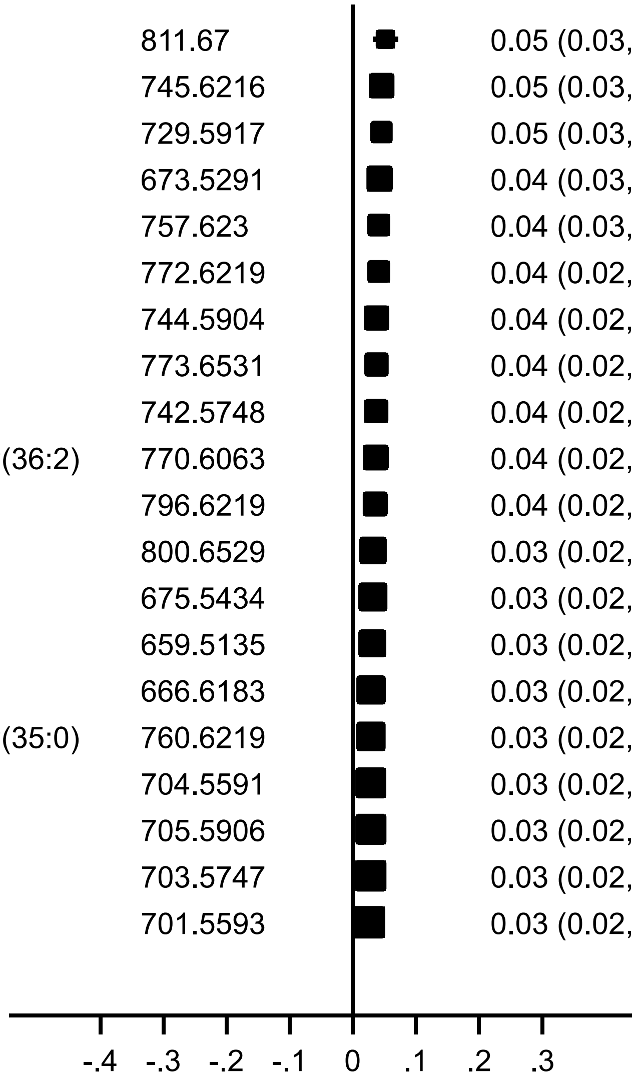

Beta (95% CI) for *GCKR* (rs1260326, chr2:27730940)

**(i) *LIPC***

| Lipid name | Lipid *m/z* | Beta (95% CI) | *P*-value |
|---|---|---|---|
| PE(36:5)-H- | 736.4922 | 0.21 (0.16, 0.25) | 2.73e-20*** |
| PE(38:6)-H- | 762.5079 | 0.16 (0.14, 0.18) | 1.06e-50*** |
| PE(34:2) | 716.523 | 0.15 (0.13, 0.17) | 9.25e-43*** |
| PE(36:4) | 740.5229 | 0.14 (0.12, 0.15) | 3.17e-54*** |
| PE(36:4)-H- | 738.5079 | 0.13 (0.11, 0.14) | 1.96e-53*** |
| Cer(42:11)+AcO- | 688.4947 | 0.11 (0.07, 0.16) | 1.84e-06* |
| PE(34:2)-H- | 714.5079 | 0.11 (0.09, 0.13) | 3.10e-32*** |
| PC(33:3) or PE(36:3) | 742.5386 | 0.11 (0.09, 0.13) | 1.35e-23*** |
| PE(40:7)-H- | 788.5236 | 0.09 (0.07, 0.11) | 7.09e-16*** |
| PC(35:4) or PE(38:4) | 768.5543 | 0.09 (0.07, 0.10) | 4.07e-41*** |
| PC(37:6) or PE(40:6) | 792.5537 | 0.07 (0.06, 0.09) | 8.49e-19*** |
| PC(35:5)-H- or PE(38:5)-H- | 764.5236 | 0.06 (0.05, 0.08) | 1.29e-12*** |
| PE(37:4)-H- or PC(34:4)-H- | 752.5235 | 0.06 (0.04, 0.07) | 1.09e-08** |
| PC(33:3)-H- or PE(36:3)-H- | 740.5236 | 0.05 (0.03, 0.06) | 2.02e-10*** |
| PC(33:2) or PE(36:2) | 744.5543 | 0.05 (0.04, 0.06) | 8.03e-16*** |
| Cer(44:11)+AcO- | 716.526 | 0.04 (0.02, 0.06) | 2.39e-05* |
| PE(34:1)-H- or PC(31:1)-H- | 716.5235 | 0.04 (0.02, 0.06) | 2.39e-05* |
| PA(39:1) | 745.5746 | 0.03 (0.02, 0.04) | 1.78e-07* |
| PC(35:4)-H- or PE(38:4)-H- | 766.5391 | 0.02 (0.01, 0.04) | 4.27e-05* |
| SM(38:1) | 759.6372 | -0.02 (-0.03, -0.01) | 6.99e-05* |

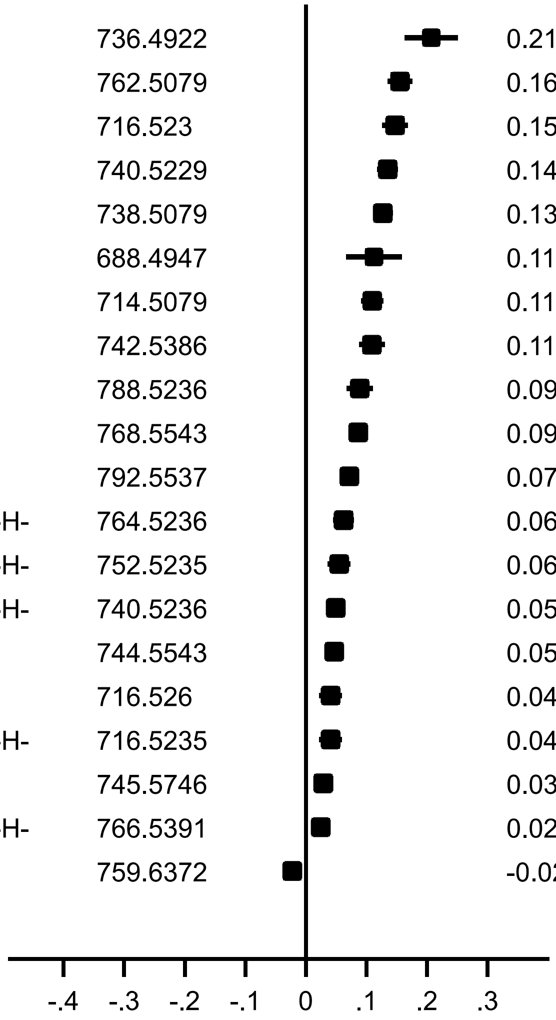


Beta (95% CI) for *LIPC* (rs2070895, chr15:58723939)

**(j)** ***LPL***

| Lipid name | Lipid *m/z* | Beta (95% CI) | *P*-value |
|---|---|---|---|
| SM(42:4) | 809.6531 | 0.08 (0.05, 0.10) | 7.51e-11*** |
| CE(20:3) | 692.6339 | 0.07 (0.04, 0.09) | 1.35e-09** |
| PC-O(39:3) or PC-P(39:2) | 812.6532 | 0.06 (0.04, 0.09) | 1.21e-08** |
| SM(42:3) | 811.6688 | 0.06 (0.04, 0.08) | 9.36e-09** |
| CE(18:0) | 670.6496 | 0.05 (0.03, 0.07) | 4.70e-09** |
| PC-O(33:1) | 732.5904 | 0.05 (0.03, 0.07) | 2.27e-08** |
| CE(18:1) | 668.6339 | 0.05 (0.03, 0.06) | 6.92e-09** |
| SM(34:1) | 703.5747 | 0.05 (0.03, 0.06) | 4.36e-08** |
| DG(36:2) | 638.5723 | -0.07 (-0.09, -0.04) | 3.85e-08** |
| DG-H20(36:2) | 603.5352 | -0.07 (-0.09, -0.05) | 3.63e-09** |
| DG-H20(34:2) | 575.5039 | -0.07 (-0.10, -0.05) | 6.57e-09** |
| DG-H20(34:1) | 577.5193 | -0.07 (-0.10, -0.05) | 3.18e-08** |
| TG(52:2) | 876.8016 | -0.08 (-0.10, -0.05) | 2.75e-08** |
| TG(53:3) | 888.8016 | -0.08 (-0.10, -0.05) | 3.63e-08** |
| TG(54:3) | 902.8175 | -0.08 (-0.10, -0.05) | 2.48e-08** |
| TG(52:3) | 874.7859 | -0.08 (-0.10, -0.05) | 2.23e-10*** |
| DG-H20(36:3) | 601.5195 | -0.08 (-0.10, -0.05) | 1.17e-08** |
| DG-H20(36:1) | 605.5508 | -0.08 (-0.11, -0.05) | 3.27e-08** |
| TG(54:4) | 900.8015 | -0.08 (-0.11, -0.05) | 3.62e-08** |
| TG(54:2) | 904.8326 | -0.09 (-0.13, -0.06) | 3.32e-08** |

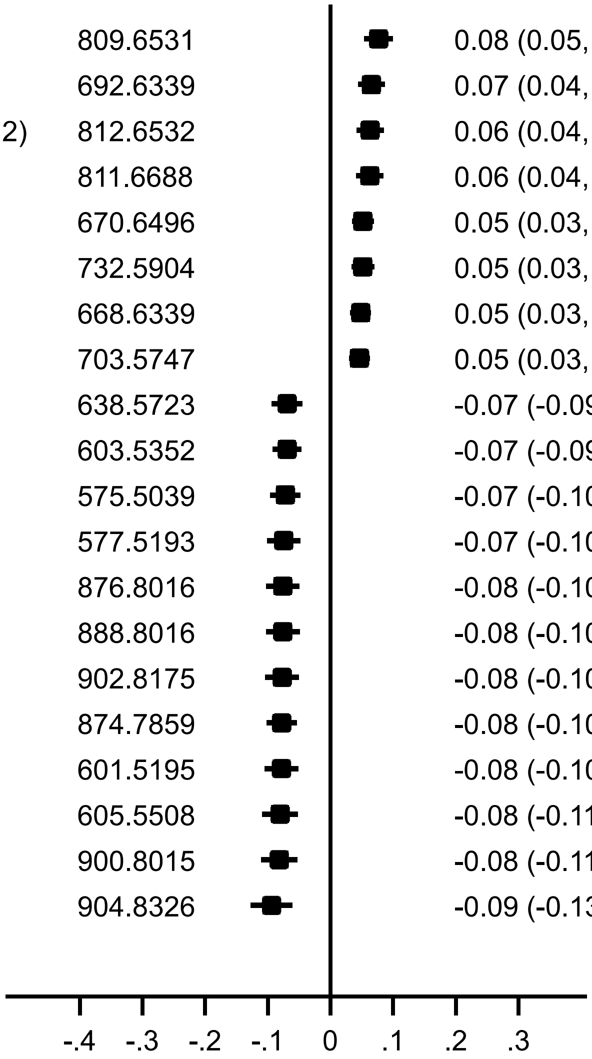


Beta (95% CI) for *LPL* (rs9644639, chr8:19884947)

#### (k) *MBOAT7*

| Lipid name | Lipid *m/z* | Beta (95% CI) | *P*-value |
|---|---|---|---|
| PI(35:2)-H- | 847.5343 | 0.12 (0.10, 0.15) | 7.88e-29*** |
| PI(32:1)-H- | 807.5028 | 0.12 (0.10, 0.15) | 1.29e-21*** |
| PI(34:0)-H- | 837.5498 | 0.12 (0.10, 0.14) | 1.08e-27*** |
| PI(40:6)-H- | 909.5498 | 0.10 (0.09, 0.12) | 1.98e-43*** |
| PC(42:11)+AcO- | 910.5603 | 0.10 (0.09, 0.12) | 8.11e-38*** |
| PC(36:6)+AcO- or PE(39:6)+AcO- | 836.5446 | 0.10 (0.08, 0.12) | 2.20e-30*** |
| PE(39:7)+AcO- or PS(40:6)-H- | 834.529 | 0.10 (0.09, 0.11) | 1.61e-45*** |
| PA(40:5)+AcO- or PG(39:5)-H- | 809.5337 | 0.10 (0.08, 0.12) | 3.31e-20*** |
| PI(34:2)-H- | 833.5186 | 0.10 (0.08, 0.11) | 8.71e-45*** |
| PI(34:1)-H- | 835.5341 | 0.09 (0.08, 0.11) | 2.56e-31*** |
| PC(38:7)+AcO- | 862.5603 | 0.09 (0.08, 0.10) | 2.85e-49*** |
| PI(36:2)-H- | 861.5498 | 0.09 (0.08, 0.10) | 6.80e-49*** |
| PI(40:5)-H- | 911.5655 | 0.08 (0.07, 0.09) | 5.26e-30*** |
| PI(38:6)-H- | 881.5186 | 0.08 (0.06, 0.09) | 2.47e-20*** |
| PI(36:1)-H- | 863.5654 | 0.06 (0.05, 0.08) | 2.47e-15*** |
| PA(44:6)+AcO- or PG(43:6)-H- | 863.5806 | 0.06 (0.05, 0.08) | 3.01e-15*** |
| PI(38:4)-H- | 885.5498 | -0.05 (-0.06, -0.04) | 5.50e-25*** |
| PC(40:9)+AcO- | 886.5603 | -0.05 (-0.07, -0.04) | 8.50e-25*** |
| PI(38:3)-H- | 887.5655 | -0.08 (-0.09, -0.06) | 2.47e-33*** |
| PC(40:8)+AcO- | 888.5759 | -0.10 (-0.11, -0.08) | 5.70e-39*** |

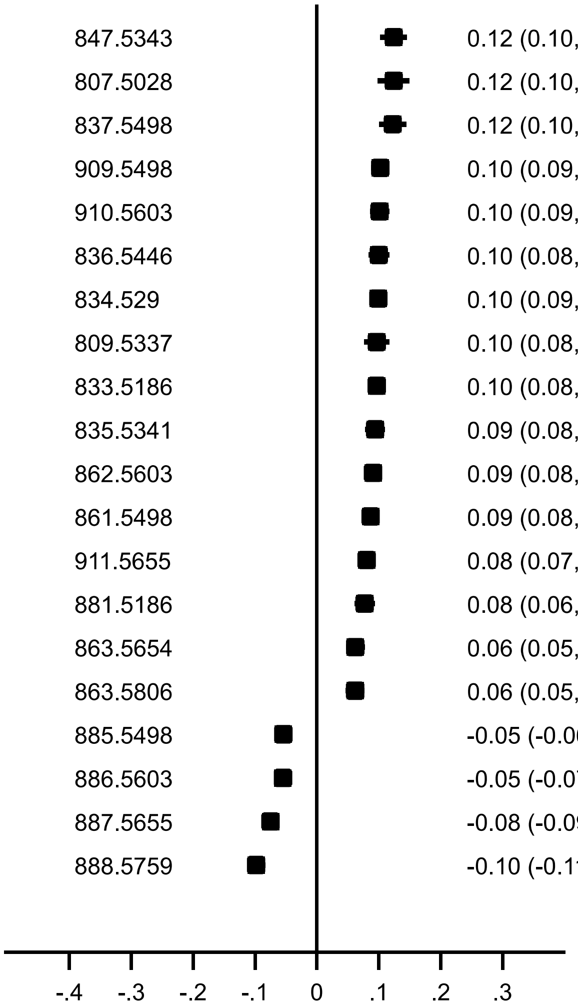


Beta (95% CI) for *MBOAT7* (rs8736, chr19:54677189)

**(I) *MLXIPL***

| Lipid name | Lipid *m/z* | Beta (95% CI) | *P*-value |
|---|---|---|---|
| PC-O(34:3) | 742.5748 | 0.06 (0.04, 0.09) | 4.52e-09** |
| SM(33:0)-H- | 689.5604 | 0.06 (0.04, 0.08) | 6.02e-10*** |
| SM(41:2)-H- | 797.6543 | 0.06 (0.04, 0.08) | 9.67e-08* |
| SM(41:1)-H- | 799.67 | 0.06 (0.04, 0.08) | 9.26e-08* |
| SM(33:1)-H- | 687.5448 | 0.06 (0.04, 0.08) | 7.76e-10*** |
| SM(42:4) | 809.6531 | 0.06 (0.03, 0.08) | 2.31e-07* |
| SM(42:3)+AcO- | 869.6754 | 0.06 (0.03, 0.08) | 6.38e-07* |
| SM(42:2)+AcO- | 871.6911 | 0.06 (0.03, 0.08) | 8.61e-07* |
| SM(34:0)+AcO- | 763.5972 | 0.05 (0.04, 0.07) | 4.02e-08** |
| SM(34:1)+AcO- | 761.5815 | 0.05 (0.03, 0.07) | 3.33e-08** |
| PC-P(34:1) | 744.5904 | 0.05 (0.03, 0.07) | 7.93e-07* |
| SM(42:0)+AcO- | 875.7224 | 0.05 (0.03, 0.07) | 1.26e-06* |
| PC-O(39:3) or PC-P(39:2) | 812.6532 | 0.05 (0.03, 0.07) | 1.14e-06* |
| SM(37:1)-H- | 743.6075 | 0.05 (0.03, 0.07) | 5.73e-07* |
| SM(42:3) | 811.6688 | 0.05 (0.03, 0.07) | 1.22e-06* |
| SM(39:2)-H- | 769.6231 | 0.05 (0.03, 0.07) | 5.29e-07* |
| SM(42:1) | 815.7001 | 0.05 (0.03, 0.06) | 2.70e-07* |
| PC-O(31:1) | 704.5591 | 0.05 (0.03, 0.06) | 3.05e-09** |
| SM(34:0) | 705.5906 | 0.05 (0.03, 0.06) | 5.83e-09** |
| SM(34:1) | 703.5747 | 0.04 (0.03, 0.06) | 6.48e-09** |

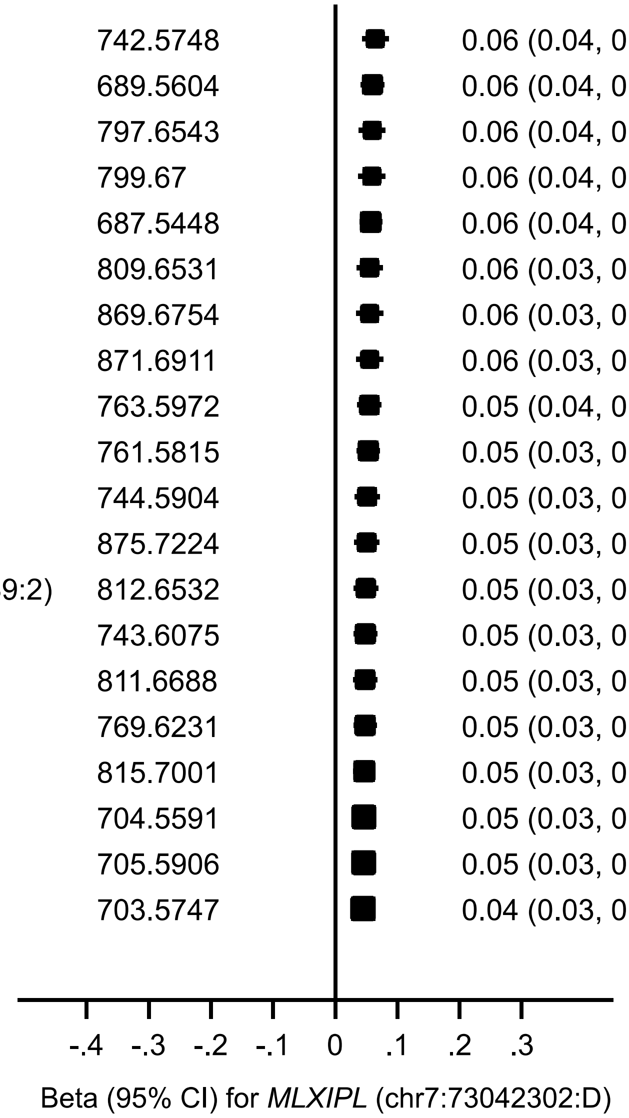

#### (m) *PAQR9*

| Lipid name | Lipid *m/z* | Beta (95% CI) | *P*-value |
|---|---|---|---|
| PA(44:6)+AcO- or PG(43:6)-H- | 863.5806 | 0.08 (0.05, 0.10) | 3.71e-10*** |
| PI(36:1)-H- | 863.5654 | 0.08 (0.05, 0.10) | 3.94e-10*** |
| PI(34:0)-H- | 837.5498 | 0.06 (0.03, 0.10) | 6.81e-04* |
| PC(36:6)+AcO- or PE(39:6)+AcO- | 836.5446 | 0.05 (0.02, 0.08) | 3.57e-04* |
| PI(34:1)-H- | 835.5341 | 0.05 (0.02, 0.07) | 2.59e-04* |
| CE(18:0) | 670.6496 | 0.02 (0.01, 0.04) | 3.07e-03 |
| CE(18:1) | 668.6339 | 0.02 (0.01, 0.04) | 3.36e-03 |
| PC(35:4)-H- or PE(38:4)-H- | 766.5391 | -0.02 (-0.03, -0.00) | 2.16e-02 |
| PA(40:2) | 757.5744 | -0.02 (-0.04, -0.00) | 2.40e-02 |
| PC(34:2)+AcO- or PS(38:1)-H- | 816.5759 | -0.02 (-0.04, -0.00) | 1.55e-02 |
| PA(45:6)-H- | 817.5752 | -0.02 (-0.04, -0.00) | 1.58e-02 |
| PS(42:8)-H- | 858.5291 | -0.03 (-0.05, -0.01) | 1.48e-02 |
| PC(35:5)-H- or PE(38:5)-H- | 764.5236 | -0.03 (-0.05, -0.00) | 2.04e-02 |
| PC(33:3)-H- or PE(36:3)-H- | 740.5236 | -0.03 (-0.05, -0.01) | 6.84e-03 |
| PE(34:2)-H- | 714.5079 | -0.03 (-0.05, -0.01) | 1.67e-02 |
| PE(36:4)-H- | 738.5079 | -0.03 (-0.06, -0.01) | 3.42e-03 |
| PE(40:7)-H- | 788.5236 | -0.03 (-0.06, -0.01) | 2.16e-02 |
| PC(34:3)+AcO- | 814.5603 | -0.04 (-0.06, -0.01) | 8.84e-04* |
| PS(40:5)+AcO- | 896.5659 | -0.05 (-0.08, -0.01) | 8.69e-03 |
| PE(36:5)-H- | 736.4922 | -0.09 (-0.15, -0.03) | 5.49e-03 |

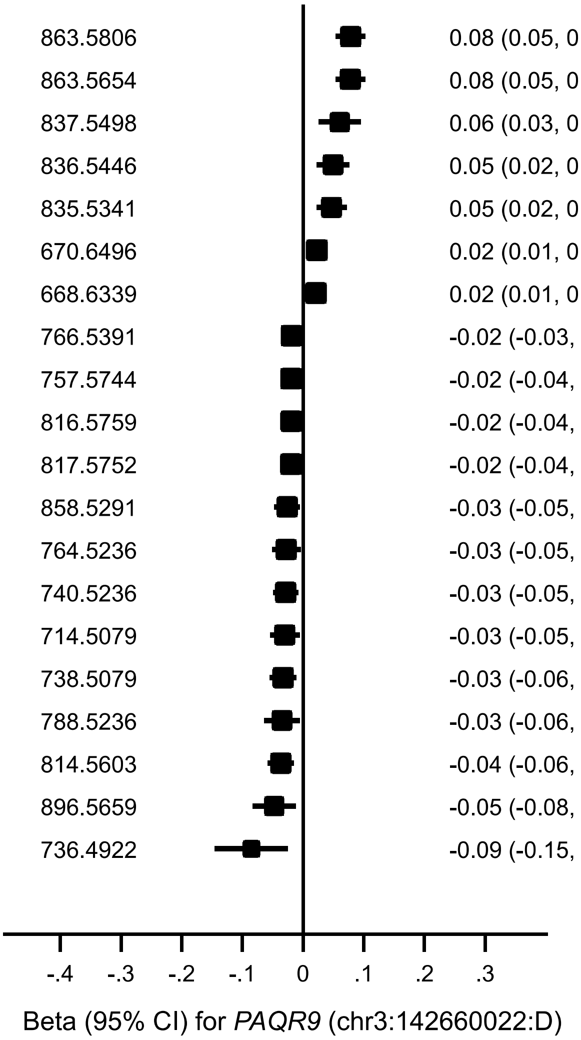

#### (n) *PCTP*

| Lipid name | Lipid *m/z* | Beta (95% CI) | *P*-value |
|---|---|---|---|
| PC-O(46:1) | 914.7941 | 0.04 (-0.01, 0.08) | 9.62e-02 |
| Cer(42:2)+AcO- | 706.6355 | 0.01 (-0.00, 0.03) | 9.67e-02 |
| PA(39:1) | 745.5746 | 0.01 (-0.00, 0.02) | 6.78e-02 |
| PE(37:4)-H- or PC(34:4)-H- | 752.5235 | -0.02 (-0.03, 0.00) | 7.34e-02 |
| PS(40:2)+AcO- | 902.6128 | -0.02 (-0.03, 0.00) | 6.91e-02 |
| PA(34:1)-H- | 673.4812 | -0.02 (-0.04, 0.00) | 9.71e-02 |
| FreeFA(21:0)-H- | 325.3113 | -0.02 (-0.04, 0.00) | 6.20e-02 |
| SM(36:1)-H- | 729.5917 | -0.02 (-0.03, -0.00) | 1.84e-02 |
| PA(44:6)+AcO- or PG(43:6)-H- | 863.5806 | -0.02 (-0.04, -0.01) | 6.58e-03 |
| PI(36:1)-H- | 863.5654 | -0.02 (-0.04, -0.01) | 6.45e-03 |
| PI(34:1)-H- | 835.5341 | -0.02 (-0.04, -0.01) | 4.41e-03 |
| PC(36:6)+AcO- or PE(39:6)+AcO- | 836.5446 | -0.02 (-0.04, -0.01) | 7.41e-03 |
| PS(41:5)+AcO- | 910.5816 | -0.03 (-0.05, 0.00) | 5.64e-02 |
| PI(32:1)-H- | 807.5028 | -0.03 (-0.06, -0.00) | 3.45e-02 |
| PI(34:0)-H- | 837.5498 | -0.03 (-0.05, -0.01) | 1.45e-02 |
| PG(34:0)+AcO- | 809.5548 | -0.04 (-0.07, -0.00) | 3.78e-02 |
| PG(32:1)+AcO- | 779.5078 | -0.04 (-0.08, -0.01) | 1.20e-02 |
| PA(40:5)+AcO- or PG(39:5)-H- | 809.5337 | -0.07 (-0.09, -0.05) | 1.98e-10*** |
| PI(33:0)-H- | 823.5341 | -0.08 (-0.15, -0.02) | 1.51e-02 |
| PI(42:8)-H- | 933.5498 | -0.09 (-0.18, 0.00) | 5.08e-02 |

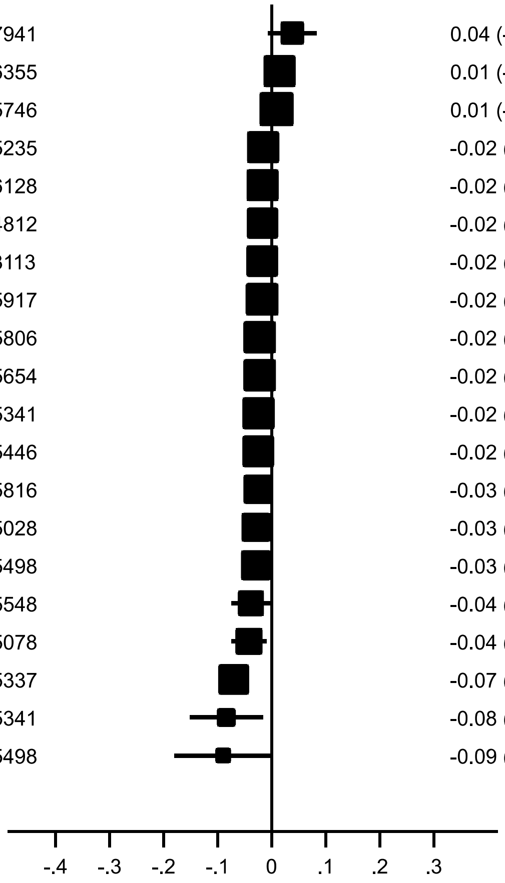


Beta (95% CI) for *PCTP* (rs11079173, chr17:53487664)

#### (o) ***PIGH-TMEM229B***

| Lipid name | Lipid *m/z* | Beta (95% CI) | *P*-value |
|---|---|---|---|
| PC(34:5)+AcO- or PE(37:5)+AcO- | 810.529 | 0.02 (-0.00, 0.05) | 5.98e-02 |
| PS(44:9)-H- | 884.5448 | 0.02 (0.00, 0.03) | 1.09e-02 |
| PC-O(32:0) | 720.5906 | 0.02 (0.00, 0.03) | 8.36e-03 |
| DG(34:2) | 610.541 | 0.02 (0.00, 0.03) | 3.37e-02 |
| PA(34:2)-H- | 671.4656 | 0.02 (0.00, 0.03) | 3.92e-02 |
| PI(38:5)-H- | 883.5343 | 0.02 (0.00, 0.03) | 3.05e-02 |
| DG(34:1) | 612.5564 | 0.02 (-0.00, 0.03) | 6.38e-02 |
| PC-P(38:3) | 796.6219 | 0.01 (0.00, 0.03) | 2.75e-02 |
| PC(40:9)+AcO- | 886.5603 | 0.01 (-0.00, 0.02) | 5.70e-02 |
| PI(38:4)-H- | 885.5498 | 0.01 (-0.00, 0.02) | 6.50e-02 |
| PC-P(33:1) | 730.5747 | -0.01 (-0.03, 0.00) | 8.49e-02 |
| SM(38:0) | 761.6532 | -0.02 (-0.03, -0.01) | 1.95e-03 |
| PA(38:1) | 731.5587 | -0.02 (-0.05, 0.00) | 7.40e-02 |
| PC-O(34:3) | 742.5748 | -0.03 (-0.04, -0.01) | 1.69e-04* |
| PS(40:1)+AcO- | 904.6284 | -0.04 (-0.07, -0.01) | 9.54e-03 |
| PG(37:0)+AcO- | 851.6017 | -0.05 (-0.06, -0.03) | 1.41e-11*** |
| PC-O(40:9) or PC-P(40:8) | 814.5744 | -0.06 (-0.11, -0.02) | 9.27e-03 |
| PC-O(36:5) | 766.5745 | -0.07 (-0.08, -0.06) | 6.45e-42*** |
| PA(41:4) | 767.559 | -0.07 (-0.08, -0.06) | 7.54e-41*** |
| SM(44:9)-H- | 825.5918 | -0.08 (-0.09, -0.07) | 4.98e-35*** |

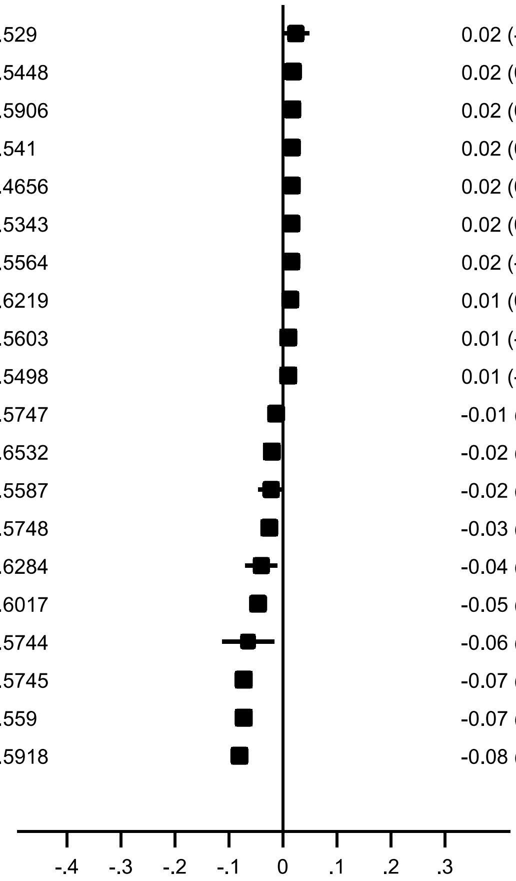


Beta (95% CI) for *PIGH-TMEM229B* (rs1885041, chr14:67976325)

#### (p) *PLA2G10-NTAN1-NPIPA5*

| Lipid name | Lipid *m/z* | Beta (95% CI) | *P*-value |
|---|---|---|---|
| PA(39:1) | 745.5746 | 0.02 (0.01, 0.03) | 3.18e-05* |
| PE(41:4) | 810.6012 | -0.02 (-0.03, -0.01) | 1.96e-04* |
| PA(42:2) | 785.6059 | -0.02 (-0.03, -0.02) | 2.44e-11*** |
| PE(39:3) | 784.5855 | -0.02 (-0.03, -0.02) | 1.85e-11*** |
| PC(36:3)+AcO- | 842.5916 | -0.02 (-0.03, -0.01) | 1.28e-06* |
| PC(35:3)-H- or PE(38:3)-H- | 768.5548 | -0.03 (-0.04, -0.02) | 4.33e-08** |
| PC(37:4)+AcO- or PE(40:4)+AcO- | 854.5915 | -0.03 (-0.04, -0.01) | 1.76e-04* |
| FreeFA(20:3)-H- | 305.2487 | -0.03 (-0.04, -0.02) | 1.98e-06* |
| PI(38:3)-H- | 887.5655 | -0.03 (-0.04, -0.02) | 3.70e-07* |
| PC(37:3) or PE(40:3) | 798.6012 | -0.03 (-0.05, -0.02) | 1.61e-07* |
| CE(20:3) | 692.6339 | -0.04 (-0.05, -0.02) | 7.47e-10*** |
| PE(40:3)+AcO- or PC(37:3)+AcO- | 856.6072 | -0.05 (-0.06, -0.03) | 2.02e-10*** |
| PC(40:8)+AcO- | 888.5759 | -0.05 (-0.06, -0.03) | 2.31e-10*** |
| PI(38:0)+AcO- | 953.6335 | -0.05 (-0.07, -0.03) | 1.86e-06* |
| PE(41:3) | 812.6168 | -0.05 (-0.06, -0.04) | 5.53e-24*** |
| PE(40:3)-H- or PC(37:3)-H- | 796.5861 | -0.05 (-0.07, -0.04) | 1.10e-17*** |
| PC(38:3)+AcO- or PE(41:3)+AcO- | 870.6229 | -0.06 (-0.07, -0.04) | 3.92e-16*** |
| PE(41:2) | 814.6322 | -0.06 (-0.07, -0.05) | 6.75e-23*** |
| PE(40:2)-H- | 798.6017 | -0.06 (-0.08, -0.05) | 7.22e-14*** |
| PE(41:2)+AcO- or PS(42:1)-H- | 872.6384 | -0.07 (-0.08, -0.05) | 5.44e-17*** |

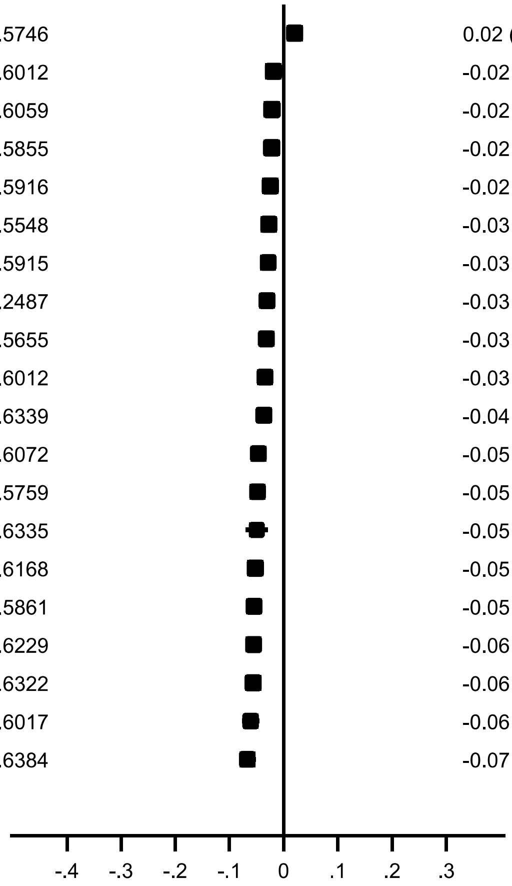


Beta (95% CI) for *PLA2G10-NTAN1-NPIPA5* (rs34955778, chr16:15139594)

#### (q) ***PNPLA3***

| Lipid name | Lipid *m/z* | Beta (95% CI) | *P*-value |
|---|---|---|---|
| TG(57:10) | 930.754 | 0.16 (0.09, 0.23) | 2.74e-06* |
| TG(58:9) | 946.7854 | 0.08 (0.04, 0.13) | 6.93e-05* |
| TG(56:6) | 924.801 | 0.07 (0.05, 0.09) | 3.37e-14*** |
| TG(55:8) | 906.7543 | 0.07 (0.03, 0.10) | 9.66e-05* |
| TG(56:5) | 926.817 | 0.07 (0.05, 0.08) | 5.85e-14*** |
| TG(56:4) | 928.8329 | 0.06 (0.04, 0.08) | 3.89e-07* |
| TG(56:7) | 922.7853 | 0.06 (0.03, 0.08) | 4.56e-07* |
| PC(40:8)+AcO- | 888.5759 | 0.04 (0.02, 0.06) | 2.31e-05* |
| TG(54:4) | 900.8015 | 0.04 (0.02, 0.06) | 1.53e-04* |
| PI(36:3)-H- | 859.5343 | 0.04 (0.02, 0.05) | 6.10e-05* |
| PI(38:3)-H- | 887.5655 | 0.03 (0.02, 0.05) | 2.62e-05* |
| PC(38:8)+AcO- or PS(42:7)-H- | 860.5447 | 0.03 (0.02, 0.05) | 1.30e-04* |
| PC-O(40:6) or PC-P(40:5) | 820.6214 | 0.02 (0.01, 0.04) | 2.70e-04* |
| PA(37:0)+AcO- | 777.5649 | 0.02 (0.01, 0.03) | 3.51e-04* |
| DG-H20(34:0) | 579.5352 | -0.04 (-0.07, -0.02) | 5.78e-05* |
| TG(50:1) | 850.7859 | -0.05 (-0.08, -0.02) | 2.95e-04* |
| TG(51:1) | 864.8016 | -0.07 (-0.10, -0.03) | 1.30e-04* |
| TG(48:0) | 824.7706 | -0.09 (-0.13, -0.05) | 3.02e-05* |
| TG(52:0) | 880.8331 | -0.09 (-0.13, -0.05) | 4.25e-06* |
| TG(46:0) | 796.7393 | -0.10 (-0.15, -0.05) | 1.82e-04* |

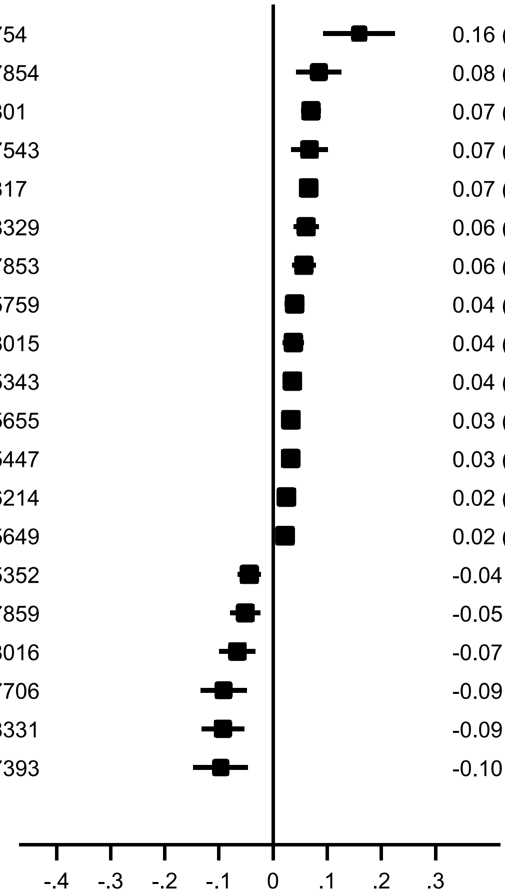


Beta (95% CI) for *PNPLA3* (rs12484809, chr22:44325631)

##### (r) *SCD*

| Lipid name | Lipid *m/z* | Beta (95% CI) | *P*-value |
|---|---|---|---|
| TG(48:0) | 824.7706 | 0.07 (0.03, 0.10) | 1.82e-04* |
| DG-H20(32:0) | 551.5038 | 0.04 (0.02, 0.06) | 6.69e-04* |
| FreeFA(16:0)-H- | 255.233 | 0.02 (0.01, 0.04) | 7.34e-04* |
| SM(36:1)-H- | 729.5917 | -0.02 (-0.04, -0.01) | 7.80e-04* |
| PG(36:2)-H- | 773.5337 | -0.03 (-0.04, -0.02) | 1.82e-05* |
| Cer(44:11)+AcO- | 716.526 | -0.03 (-0.05, -0.01) | 2.82e-04* |
| PE(34:1)-H- or PC(31:1)-H- | 716.5235 | -0.03 (-0.05, -0.01) | 2.82e-04* |
| PS(38:1)+AcO- | 876.5971 | -0.03 (-0.05, -0.01) | 6.39e-04* |
| TG(50:3) | 846.7546 | -0.03 (-0.05, -0.02) | 2.42e-05* |
| PC(40:7)+AcO- | 890.5916 | -0.03 (-0.05, -0.02) | 1.28e-06* |
| DG-H20(34:3) | 573.488 | -0.04 (-0.05, -0.02) | 2.66e-05* |
| PC(32:1) | 732.5541 | -0.04 (-0.05, -0.02) | 1.22e-04* |
| PE(40:7)-H- | 788.5236 | -0.04 (-0.05, -0.02) | 1.62e-04* |
| PI(38:1)-H- | 891.5967 | -0.04 (-0.05, -0.02) | 1.26e-06* |
| PA(38:0) | 733.5747 | -0.04 (-0.06, -0.02) | 1.01e-04* |
| PC(32:1)+AcO- or PE(35:1)+AcO- | 790.5602 | -0.04 (-0.06, -0.02) | 5.81e-05* |
| LysoPC(16:1) | 494.3245 | -0.05 (-0.06, -0.03) | 3.60e-11*** |
| CE(16:1) | 640.6024 | -0.05 (-0.07, -0.03) | 2.46e-08** |
| FreeFA(16:1)-H- | 253.2174 | -0.07 (-0.10, -0.05) | 5.80e-10*** |
| Cer(42:11)+AcO- | 688.4947 | -0.08 (-0.12, -0.04) | 4.20e-05* |

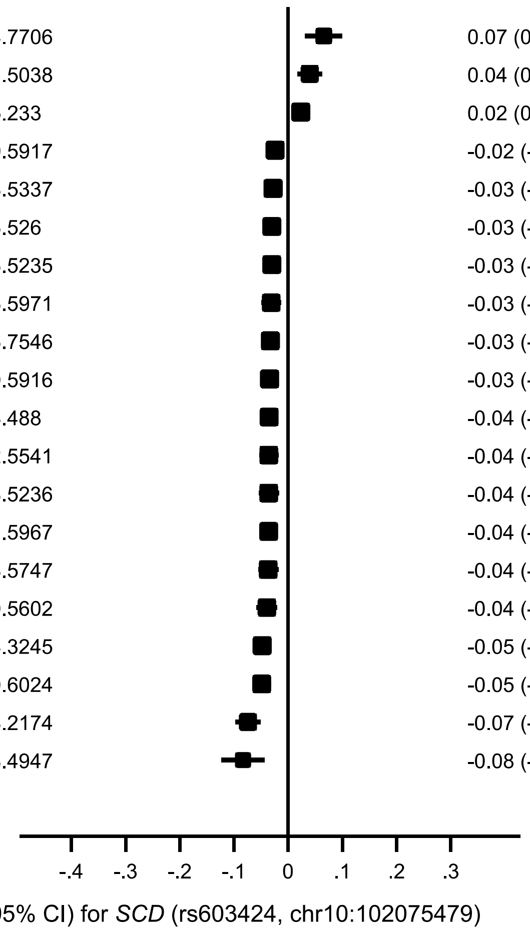

#### (s) *SGPP1*

| Lipid name | Lipid *m/z* | Beta (95% CI) | *P*-value |
|---|---|---|---|
| SM(32:1)+AcO- | 733.5502 | 0.09 (0.07, 0.10) | 1.75e-26*** |
| SM(32:1) | 675.5434 | 0.09 (0.07, 0.10) | 8.21e-35*** |
| SM(31:1)-H- | 659.5135 | 0.08 (0.06, 0.09) | 4.31e-25*** |
| PC-O(36:1) or PC-P(36:0) | 774.6376 | 0.06 (0.03, 0.09) | 1.92e-05* |
| SM(39:1) | 773.6531 | 0.06 (0.04, 0.07) | 3.41e-10*** |
| SM(38:1)-H- | 757.623 | 0.05 (0.03, 0.07) | 6.21e-08* |
| SM(39:1)+AcO- | 831.6597 | 0.05 (0.03, 0.07) | 1.22e-06* |
| SM(38:0) | 761.6532 | 0.05 (0.03, 0.07) | 7.33e-08* |
| SM(32:1)-H- | 673.5291 | 0.05 (0.03, 0.06) | 8.15e-09** |
| SM(33:0)+AcO- | 749.5815 | 0.04 (0.02, 0.06) | 1.26e-04* |
| SM(39:2)+AcO- | 829.6442 | 0.04 (0.02, 0.06) | 1.53e-04* |
| SM(37:1)+AcO- | 803.6286 | 0.04 (0.02, 0.06) | 6.00e-05* |
| SM(38:0)+AcO- | 819.6598 | 0.04 (0.02, 0.06) | 5.80e-05* |
| PC-O(35:1) or PC-P(35:0) | 760.6219 | 0.04 (0.02, 0.05) | 3.58e-07* |
| SM(38:1)+AcO- | 817.6441 | 0.04 (0.02, 0.05) | 1.56e-05* |
| SM(38:1) | 759.6372 | 0.03 (0.02, 0.05) | 5.09e-07* |
| SM(37:1) | 745.6216 | 0.03 (0.02, 0.05) | 1.15e-04* |
| SM(37:1)-H- | 743.6075 | 0.03 (0.02, 0.05) | 1.46e-04* |
| SM(40:2) | 785.6529 | 0.03 (0.01, 0.04) | 3.78e-05* |
| PC-P(37:1) | 786.6373 | 0.03 (0.01, 0.04) | 1.50e-04* |

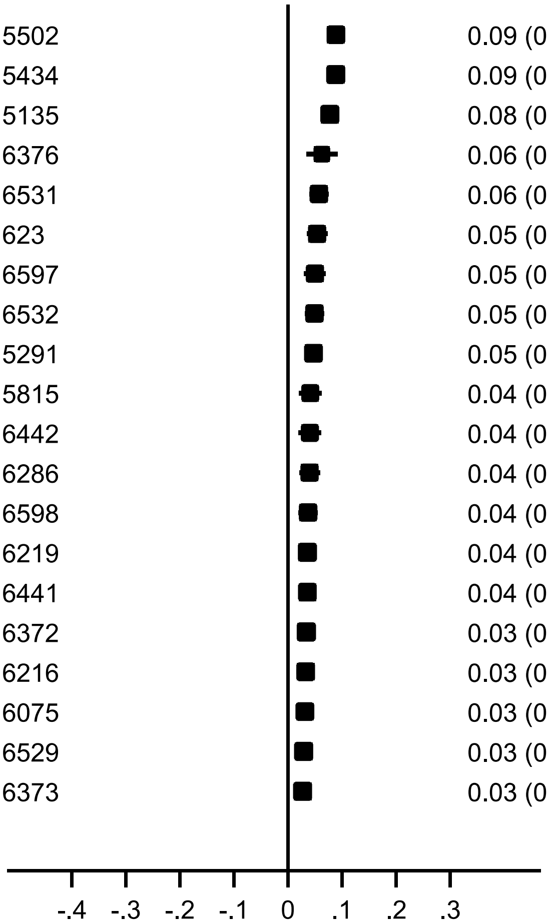


Beta (95% CI) for *SGPP1* (rs7157785, chr14:64235556)

**(t) *SPTLC3***

| Lipid name | Lipid *m/z* | Beta (95% CI) | *P*-value |
|---|---|---|---|
| PC-O(35:1) or PC-P(35:0) | 760.6219 | -0.04 (-0.06, -0.03) | 4.68e-16*** |
| SM(38:1) | 759.6372 | -0.05 (-0.06, -0.03) | 1.95e-17*** |
| SM(32:1) | 675.5434 | -0.05 (-0.06, -0.04) | 2.67e-20*** |
| SM(32:1)+AcO- | 733.5502 | -0.05 (-0.07, -0.04) | 4.34e-16*** |
| Cer(42:1)-H- | 648.63 | -0.07 (-0.08, -0.05) | 6.13e-15*** |
| SM(39:1)+AcO- | 831.6597 | -0.07 (-0.08, -0.05) | 9.54e-18*** |
| SM(37:1)+AcO- | 803.6286 | -0.07 (-0.09, -0.06) | 3.32e-20*** |
| SM(37:1) | 745.6216 | -0.07 (-0.09, -0.06) | 4.02e-29*** |
| SM(36:1)-H- | 729.5917 | -0.07 (-0.09, -0.06) | 3.67e-21*** |
| SM(38:1)-H- | 757.623 | -0.08 (-0.09, -0.06) | 1.71e-22*** |
| Cer(40:1)-H- | 620.5987 | -0.08 (-0.10, -0.06) | 9.77e-20*** |
| SM(39:1) | 773.6531 | -0.08 (-0.10, -0.07) | 1.66e-30*** |
| Cer(42:0)-H- | 650.6457 | -0.08 (-0.10, -0.06) | 8.19e-17*** |
| Cer(41:1)-H- | 634.6144 | -0.09 (-0.11, -0.07) | 2.09e-17*** |
| SM(42:2)-H- | 811.67 | -0.09 (-0.11, -0.07) | 1.09e-20*** |
| Cer(40:2)-H- | 618.5831 | -0.09 (-0.12, -0.07) | 1.31e-16*** |
| Cer(41:2)-H- | 632.5987 | -0.11 (-0.13, -0.08) | 2.81e-18*** |
| PC-O(36:1) or PC-P(36:0) | 774.6376 | -0.11 (-0.13, -0.09) | 1.80e-21*** |
| Cer(40:0)-H- | 622.6144 | -0.12 (-0.14, -0.09) | 7.97e-20*** |
| Cer(41:0)-H- | 636.6301 | -0.13 (-0.16, -0.10) | 1.95e-17*** |

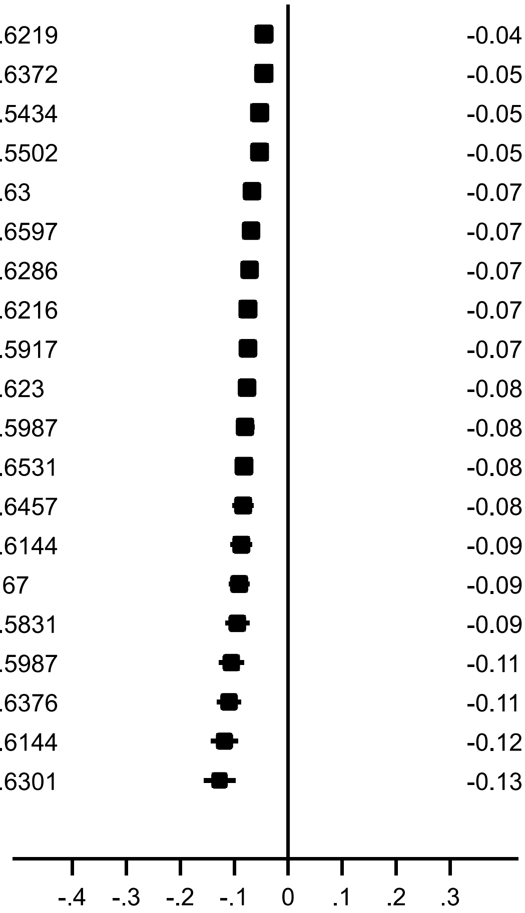


Beta (95% CI) for *SPTLC3* (rs438568, chr20:12958687)

#### (u) *UGT8*

| Lipid name | Lipid *m/z* | Beta (95% CI) | *P*-value |
|---|---|---|---|
| PG(32:1)+AcO- | 779.5078 | 0.13 (0.10, 0.17) | 4.60e-14*** |
| PI(42:8)-H- | 933.5498 | 0.10 (0.00, 0.19) | 4.00e-02 |
| PS(40:1)+AcO- | 904.6284 | 0.04 (0.01, 0.07) | 1.67e-02 |
| PS(38:4)+AcO- | 870.5502 | 0.04 (0.00, 0.08) | 4.61e-02 |
| PS(36:1)+AcO- | 848.5658 | 0.03 (0.01, 0.05) | 1.57e-02 |
| PC(33:3)+AcO- or PE(36:3)+AcO- | 800.5446 | 0.02 (-0.00, 0.05) | 8.18e-02 |
| LysoPC(18:0) | 524.3717 | 0.01 (-0.00, 0.03) | 5.27e-02 |
| LysoPC(18:1) | 522.356 | 0.01 (-0.00, 0.03) | 6.12e-02 |
| PE(41:3) | 812.6168 | 0.01 (0.00, 0.02) | 4.66e-02 |
| PC(34:2) | 758.57 | -0.01 (-0.02, -0.00) | 2.91e-02 |
| SM(36:1)+AcO- | 789.6128 | -0.01 (-0.02, 0.00) | 8.61e-02 |
| SM(34:1)+AcO- | 761.5815 | -0.01 (-0.02, -0.00) | 5.15e-02 |
| SM(32:1)+AcO- | 733.5502 | -0.01 (-0.03, 0.00) | 5.55e-02 |
| PC(34:2)+AcO- or PS(38:1)-H- | 816.5759 | -0.01 (-0.02, -0.00) | 2.02e-02 |
| SM(34:0)+AcO- | 763.5972 | -0.01 (-0.03, -0.00) | 4.13e-02 |
| PA(45:6)-H- | 817.5752 | -0.01 (-0.02, -0.00) | 1.90e-02 |
| SM(40:0)+AcO- | 847.6911 | -0.01 (-0.03, -0.00) | 4.15e-02 |
| SM(40:3)+AcO- | 841.6442 | -0.02 (-0.03, 0.00) | 6.64e-02 |
| SM(40:7) | 775.5743 | -0.08 (-0.16, -0.00) | 4.50e-02 |
| SM(42:9) | 799.5743 | -0.10 (-0.19, -0.02) | 1.59e-02 |

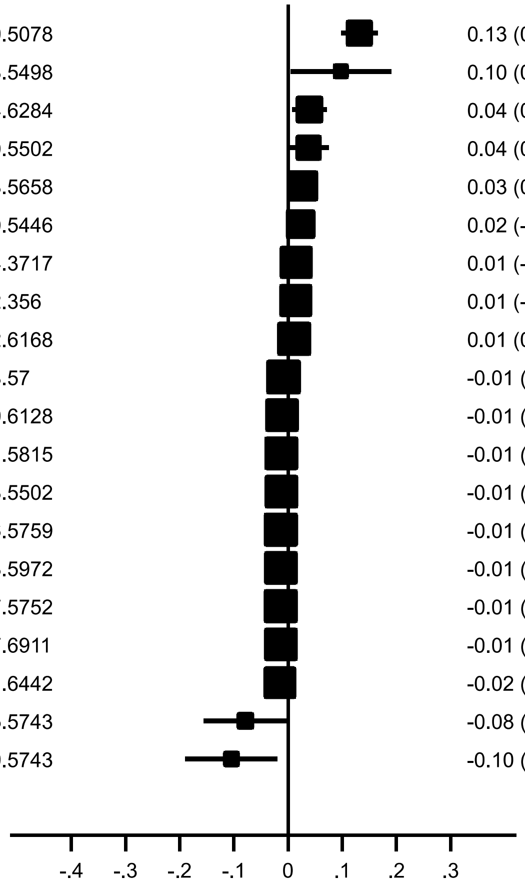


Beta (95% CI) for *UGT8* (rs28870381, chr4:115478499)

#### (v) *XBP1*

| Lipid name | Lipid *m/z* | Beta (95% CI) | *P*-value |
|---|---|---|---|
| PC-O(39:1) or PC-P(39:0) | 816.6845 | -0.09 (-0.14, -0.04) | 1.10e-04* |
| SM(36:1) | 731.6062 | -0.10 (-0.15, -0.05) | 8.79e-05* |
| SM(34:1) | 703.5747 | -0.10 (-0.15, -0.05) | 1.79e-05* |
| SM(32:1) | 675.5434 | -0.11 (-0.16, -0.06) | 2.68e-05* |
| PC-O(31:1) | 704.5591 | -0.11 (-0.15, -0.06) | 9.78e-06* |
| SM(34:2) | 701.5593 | -0.12 (-0.17, -0.07) | 6.63e-06* |
| SM(40:2) | 785.6529 | -0.12 (-0.18, -0.07) | 6.41e-06* |
| SM(41:2) | 799.6685 | -0.13 (-0.19, -0.07) | 1.17e-05* |
| SM(40:1) | 787.6688 | -0.13 (-0.18, -0.08) | 1.18e-07* |
| SM(34:0) | 705.5906 | -0.13 (-0.18, -0.08) | 5.05e-07* |
| SM(41:1) | 801.6844 | -0.13 (-0.19, -0.07) | 1.03e-05* |
| SM(42:1) | 815.7001 | -0.14 (-0.20, -0.09) | 3.86e-07* |
| PC-O(36:3) or PC-P(36:2) | 770.6063 | -0.15 (-0.21, -0.08) | 7.02e-06* |
| PC(34:2)-H- | 756.5548 | -0.15 (-0.23, -0.07) | 1.31e-04* |
| PC-O(37:1) | 788.653 | -0.16 (-0.21, -0.11) | 3.21e-09** |
| PE(38:1) | 774.6009 | -0.17 (-0.24, -0.10) | 1.19e-06* |
| SM(39:1) | 773.6531 | -0.18 (-0.24, -0.11) | 7.14e-08* |
| PC(35:2) | 772.5856 | -0.18 (-0.25, -0.12) | 5.71e-08* |
| LysoPC(17:0) | 510.356 | -0.22 (-0.33, -0.12) | 3.26e-05* |
| SM(37:1) | 745.6216 | -0.23 (-0.30, -0.16) | 2.67e-11*** |

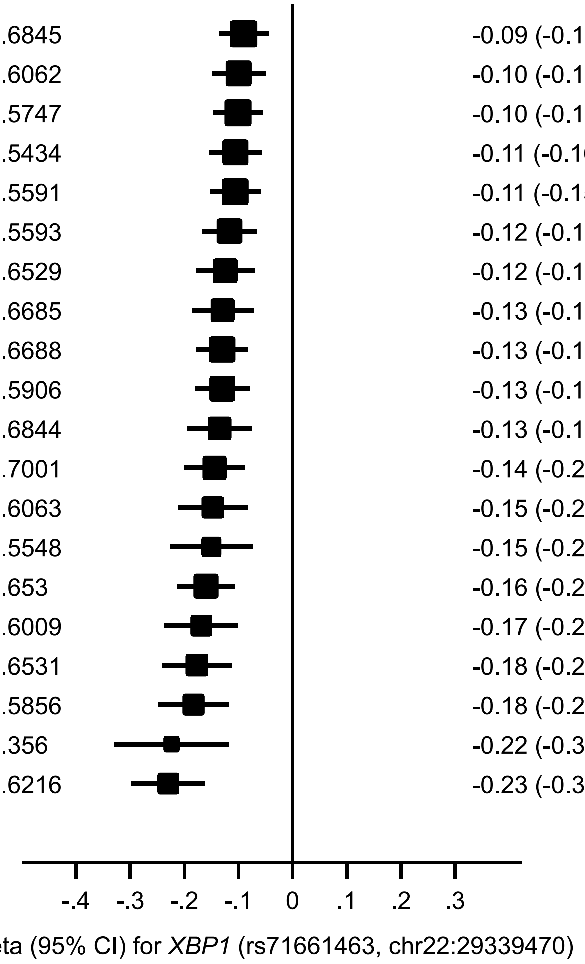


Forest plots showing the association of the top 20 most significantly associated lipids with the lead variant in each significant locus from the conditional analyses. **Note:** * = $P < 0.001$; ** = $P < 5 \times 10^{-8}$; *** = $P < 8.9 \times 10^{-10}$.
