## Supplementary figures and images for "Genome-wide analysis of blood lipid metabolites in over 5,000 South Asians reveals biological insights at cardiometabolic disease loci"

### Additional file 3 - High-resolution version of Supplementary Figure 1

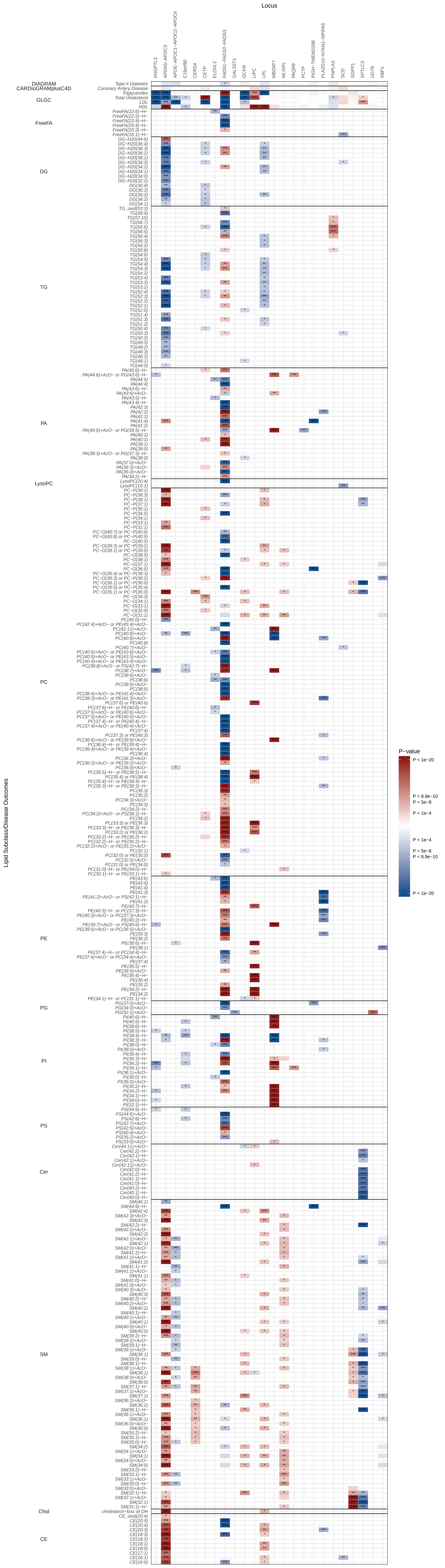
